# Morphological set enrichment enables interpretable prognostication and molecular profiling of meningiomas

**DOI:** 10.64898/2026.02.23.26346491

**Authors:** Marina A. Ayad, Kathleen McCortney, Harrshavasan T.S. Congivaram, Mateo G. Hjerthen, Alicia Steffens, Hui Zhang, Mark W. Youngblood, Amy B. Heimberger, James P. Chandler, Pouya Jamshidi, Jared T. Ahrendsen, Stephen T. Magill, David R. Raleigh, Craig M. Horbinski, Lee A.D. Cooper

## Abstract

Meningiomas are the most common primary brain tumors and, despite their benign reputation, often behave aggressively. Meningiomas are morphologically heterogeneous, yet the full significance of their histologic diversity is unclear. This is in large part because many features are not readily quantifiable by traditional observer-based light microscopy. Molecular testing improves prognostic stratification, but is not universally accessible. We therefore sought to determine whether an artificial intelligence (AI)-trained program could predict specific genomic and epigenomic patterns in meningiomas, and whether it could extract more prognostic information out of standard hematoxylin and eosin (H&E) histopathology than the current WHO classification. To do this, we developed Morphologic Set Enrichment (MSE), an interpretable computational pathology framework that quantifies statistical enrichment of morphologic patterns, cells, and tissue architecture from H&E whole-slide images. The MSE meningioma histology program was able to accurately predict DNA methylation subtypes and concurrent chromosome 1p/22q losses, in the process identifying specific morphologic patterns associated with key genomic and epigenomic alterations. It also added prognostic value independent of standard clinical and pathological variables. These results demonstrate that AI-based quantitative morphologic profiling can capture clinically and biologically relevant information that redefines risk stratification for meningiomas, incorporating histological information not included in existing grading schemes.

## INTRODUCTION

Meningiomas are the most common primary intracranial tumor [1], with over 37,000 new cases diagnosed annually in the United States [2–4]. They arise from the meningeal surfaces of the brain and spinal cord [5–7]. Known risk factors include advanced age, male sex, African descent, ionizing radiation exposure, and certain familial genetic diseases. Although commonly thought of as benign, many meningiomas are more aggressive, repeatedly recurring and invading into the underlying brain. In fact, a recent study indicated that meningioma patients have 14-20 average years of life lost, almost as much as glioblastoma patients [8].

The World Health Organization (WHO) grading of meningiomas is based on histology and certain molecular features [7]. Approximately 80% of meningiomas are CNS WHO grade 1, with <2.5 mitoses/mm^2^ and no brain invasion. Grade 2 meningiomas (18%) are intermediate risk and are defined by either a mitotic rate of 2.5-11 mitoses/mm^2^, brain invasion, and/or 3+ other high risk histologic features. Those other high risk features include necrosis, patternless sheet-like growth, prominent nucleoli (macronucleoli), high cellularity, and high nuclear to cytoplasmic ratio. Grade 3 (2%) have ≥12.5 mitoses/mm^2^, *TERT* promoter mutation, and/or homozygous *CDKN2A/B* deletion [3]. Clinical management depends on tumor location, grade, and symptoms. Adjuvant radiotherapy is included in the standard of care for grade 3 meningiomas, while grade 2 lesions may also be irradiated [9]. However, many tumors behave either more or less aggressively than would have been predicted by histologic grade alone. This is in part because histologic assessment is highly subjective, with low interobserver concordance, and is heavily dependent on observer experience [10–12]. This is also because many histopathologic features that might have prognostic value, like whorls, are impractical for even highly skilled human observers to quantify.

To improve diagnostic and prognostic accuracy, even more molecular biomarkers are being incorporated into recommendations by The Consortium to Inform Molecular and Practical Approaches to CNS Tumor Taxonomy (c-IMPACT NOW), including chromosomal arm losses like those involving 1p/22q, mRNA expression risk score, and DNA methylation signatures [13–24]. Several methylation classifiers have also been developed to improve prognostic stratification [25–27]. While molecular screening is more objective, most patients do not have access to such advanced testing. Thus, histopathologic evaluation remains the only common modality for the workup of meningiomas. New ways of objectively quantifying morphology may therefore improve prognostication and reveal novel associations between histopathologic patterns, molecular biomarkers, and clinical outcomes. A recent examination of molecular subtyping highlights the impact of the tumor microenvironment (TME) and immune infiltration on methylation based classifiers, and suggests that risk exists on a continuum modulated by the TME rather than in discrete subsets reflecting tumor cell characteristics [28]. This study highlights the importance of examining the orthogonal dimension of the TME and the potential utility for histology.

Artificial intelligence (AI) research has been developing models to predict diagnosis, molecular biomarkers, WHO grade, and/or prognosis from histology in a variety of settings [29–34], including meningiomas [34–37]. To date, none have attempted to predict multidimensional molecular profiles and clinical outcomes using models developed exclusively for meningiomas. In the current study, we sought to develop a new comprehensive AI model of meningiomas to explore the relationships between morphologic patterns, prognostic DNA and molecular biomarkers, and recurrence-free survival (**Fig. 1**). This involved a novel Morphology Set Enrichment (MSE) approach that provides strong interpretability, incorporating prior knowledge of morphologic patterns from human observers without requiring tedious manual scoring of those patterns. MSE also captures the continuous and heterogenous nature of tissues by providing a quantitative measure of targeted morphologic patterns, enabling more consistent, robust, and interpretable downstream statistical analyses. Our results demonstrate the ability of AI-trained programs to extract more meaningful predictive and prognostic information from routine meningioma histopathology than human observers ever could.

**Figure 1.**
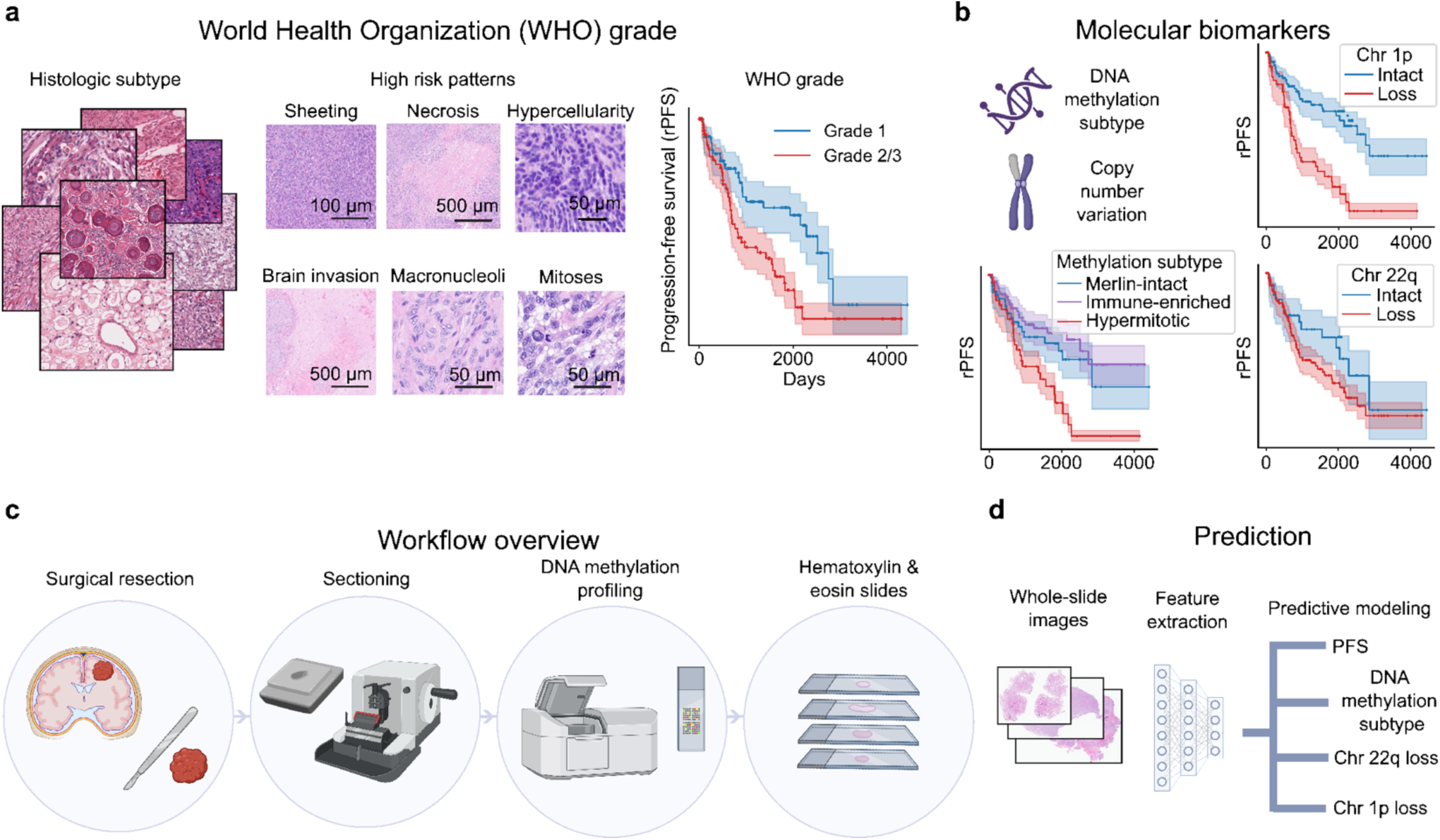
Clinical prognostication and analytic workflow for meningiomas. World Health Organization (WHO) grading assigns meningiomas to three prognostic groups based on histologic subtypes and the presence and semi-quantified high-risk morphologic patterns. **(a)** WHO grade Kaplan-Meier plots of progression free survival for the discovery cohort of 345 meningiomas. **(b)** DNA methylation subtypes have links to established high risk genetic features like the combined losses of chromosomes 1p and 22q. **(c)** Overview of the analytical workflow starting from tumor resection to hematoxylin and eosin (H&E) staining and whole-slide imaging for computational analysis. **(d)** Whole slide images of H&E-stained slides were analyzed to predict progression free survival, DNA methylation subtypes, and chromosomal losses of 1p and 22q.

## RESULTS

### Mapping morphologic patterns in meningioma

The dominant paradigm in computational pathology is Multiple Instance Learning (MIL), where models consume pixels and predict slide- or patient-level labels like diagnosis, biomarker status, or clinical outcome. MIL obviates the need for annotating tissue structures and learns directly from clinical data, and thus is popular for many applications. Learning directly from pixels avoids the bias of prior knowledge and identifies latent patterns, but may also lead to spurious correlations and overfitting [38–41]. Furthermore, while MIL models are easier to develop and often provide superior predictive accuracy compared to traditional observer-based microscopy [42–45], MIL prediction mechanisms are opaque, and their explanations for specific predictions are sometimes unsatisfactory. MIL models typically generate attention heatmaps indicating the saliency of regions for during prediction; however, heatmaps only indicate the areas where the model is focused, leaving what patterns the model perceives to the intuition of observers [46, 47].

To overcome MIL limitations, we developed Morphology Set Enrichment (MSE), a classification-free approach to identifying patterns in whole-slide images by statistical analysis of latent embeddings (**Fig. 2**). MSE was inspired by bioinformatics techniques that use the concept of statistical enrichment to identify biological themes that are overrepresented in collections of genes. We adapted this concept to histology by using an embedding space to measure the similarity between high-power fields (HPFs) from a whole slide image and a curated library of HPFs representing patterns of interest (**Fig. 2a**). By comparing these similarities to those of randomly sampled tiles (**Fig. 2b**), MSE detects whether the tiles are more similar than expected by chance using rank statistics. This method avoids the need for pattern classification models that are burdensome to develop and validate when the patterns of interest are numerous and diverse, as they are in meningiomas.

**Figure 2.**
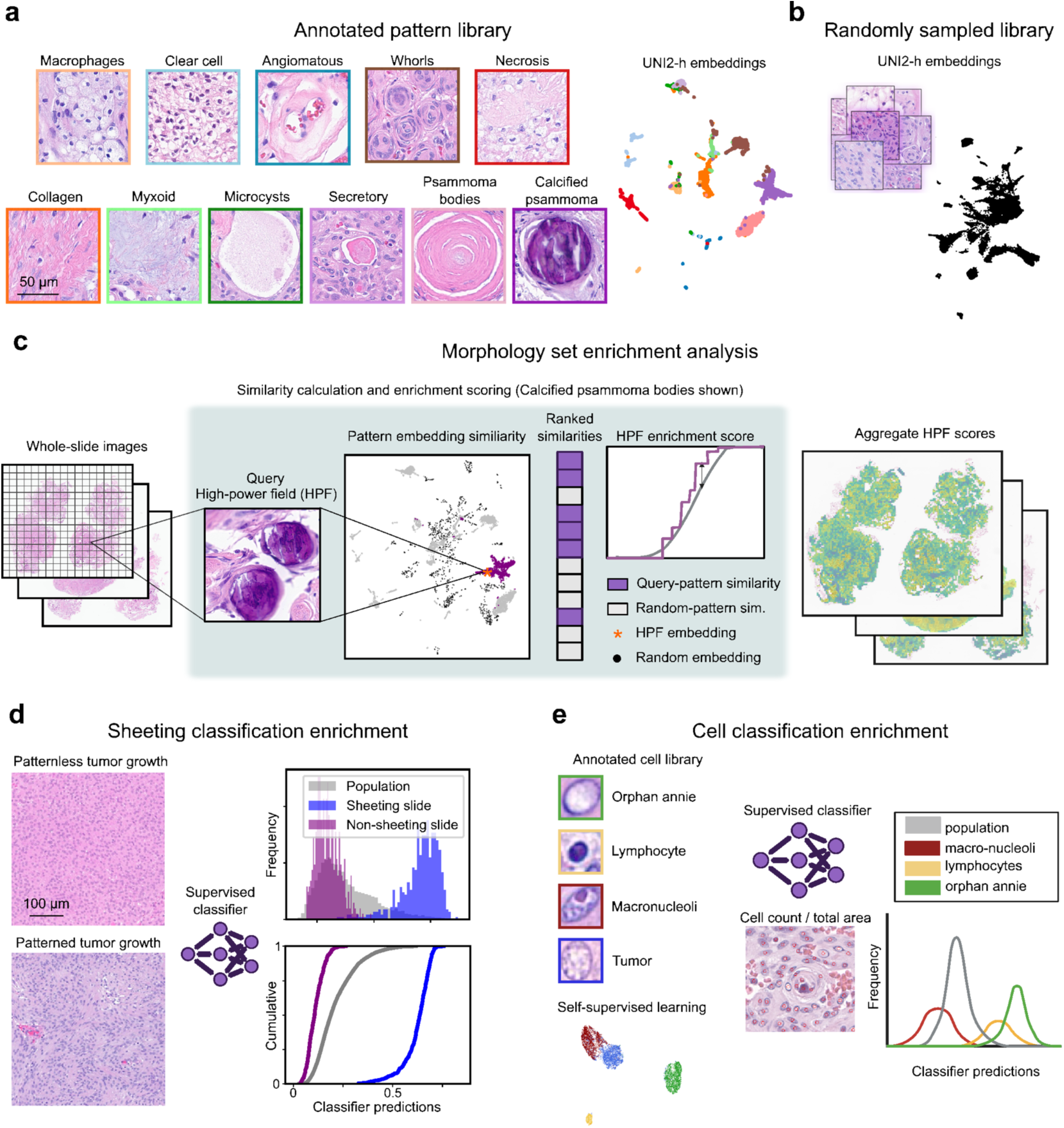
Morphology Set Enrichment (MSE) analysis overview. We developed a method to measure the enrichment of meningioma morphologic patterns using similarity statistics. **(a)** A library containing annotated examples of high-power fields (HPFs) from 11 patterns was developed from the pattern cohort. HPFs were embedded in a latent space using the UNI2-h model. **(b)** A library of randomly sampled HPFs was also generated from the pattern cohort. **(c)** MSE measures the enrichment or depletion of histology patterns in a query specimen by comparing the similarities of query HPFs and random HPFs with the pattern library. A query HPF is compared to each example of a pattern to calculate similarities in the latent space. Randomly sampled HPFs are also compared to the pattern to provide a baseline of expected similarities. A pattern enrichment score is calculated by comparing the rank statistics of the query HPF similarities and random similarities. This is repeated for each pattern and aggregated over all HPFs to generate specimen-level enrichment scores. **(d)** Enrichment of sheeting was measured by classification of low-power fields to discriminate between pattern-less and patterned tumor growth. **(e)** Cell type enrichment was calculated by nuclear classification at high-resolution. Cell features from our cell dictionary are shown with clear clustering between different cell types.

We defined 11 patterns for meningiomas and generated a library containing 100 to 1,200 examples for each pattern (**Fig. 2a**). These choices were motivated by characteristic patterns of WHO histologic subtypes, high-risk patterns [7], speculation about the trajectories of transitions between patterns associated with disease progression [48–50], and interest in immune infiltrates [51, 52]. This library comprised 12,484 HPF images from a “pattern cohort” which was distinct from the cohort used to evaluate associations between patterns, biomarkers, and clinical outcomes. HPFs were embedded using the UNI-2h model at 200× magnification, and visualization of those embeddings showed that patterns are distinguishable in the embedding space (**Fig. 2a**). Patient-level pattern enrichment was calculated by first measuring HPF-level pattern enrichments, then aggregating the median across all HPFs and slides for a patient (**Fig. 2c**). Different aggregation methods were explored (**Fig. S1**).

Patternless or “sheeting” growth is an adverse prognostic morphologic pattern that is challenging to assess, as indicated by low interobserver concordance [12]. Distinguishing subtle patterns like patterned vs. patternless tumor growth requires the incorporation of negative examples, which is not currently possible with MSE. To address this, we developed a supervised classifier of sheeting at 50× magnification (AUC 0.99, balanced accuracy 0.96). This classifier was used to calculate the abundance of patternless tumor in each patient, which was transformed into a sheeting enrichment score by comparing patternless growth abundance to cohort-wide statistics (**Fig. 2d**).

Cellular features of meningiomas, including lymphocytes, tumor nuclei with chromatin clearing (“Orphan Annie”), and macronucleoli, cannot be reliably measured from HPF embeddings due to a lack of sensitivity. These features are often not the dominant pattern within an HPF, with many HPFs containing only one or several instances. To measure these features, we developed a complementary nuclear morphology classifier by supervised learning (**Fig. 2e**). We first created a self-supervised model of single-cell nuclear morphology, using CellViT++, to segment individual cell nuclei [53]. Embeddings from this model were then used to develop a cell nuclei classifier, resulting in an AUC of 0.99 and a balanced accuracy of 0.941. Abundances of cell types in each patient were transformed to enrichment scores based on cohort-wide abundance statistics.

### Consensus and divergence between MSE and neuropathologists in evaluating meningiomas

MSE scores were compared to ordinal ratings from three board-certified neuropathologists (PJ, JTA, CMH) to compare pathologist-pathologist and machine-pathologist agreement (**Fig. 3a**). Pathologist agreement ranged from poor (clear cell, ICC=0.16) to substantial (calcified psammoma bodies, ICC=0.93). The highest machine-pathologist agreement ranged from weak (secretory, ICC=0.27) to high (psammoma bodies, ICC=0.78) and generally correlated with pathologist-pathologist agreement.

**Figure 3.**
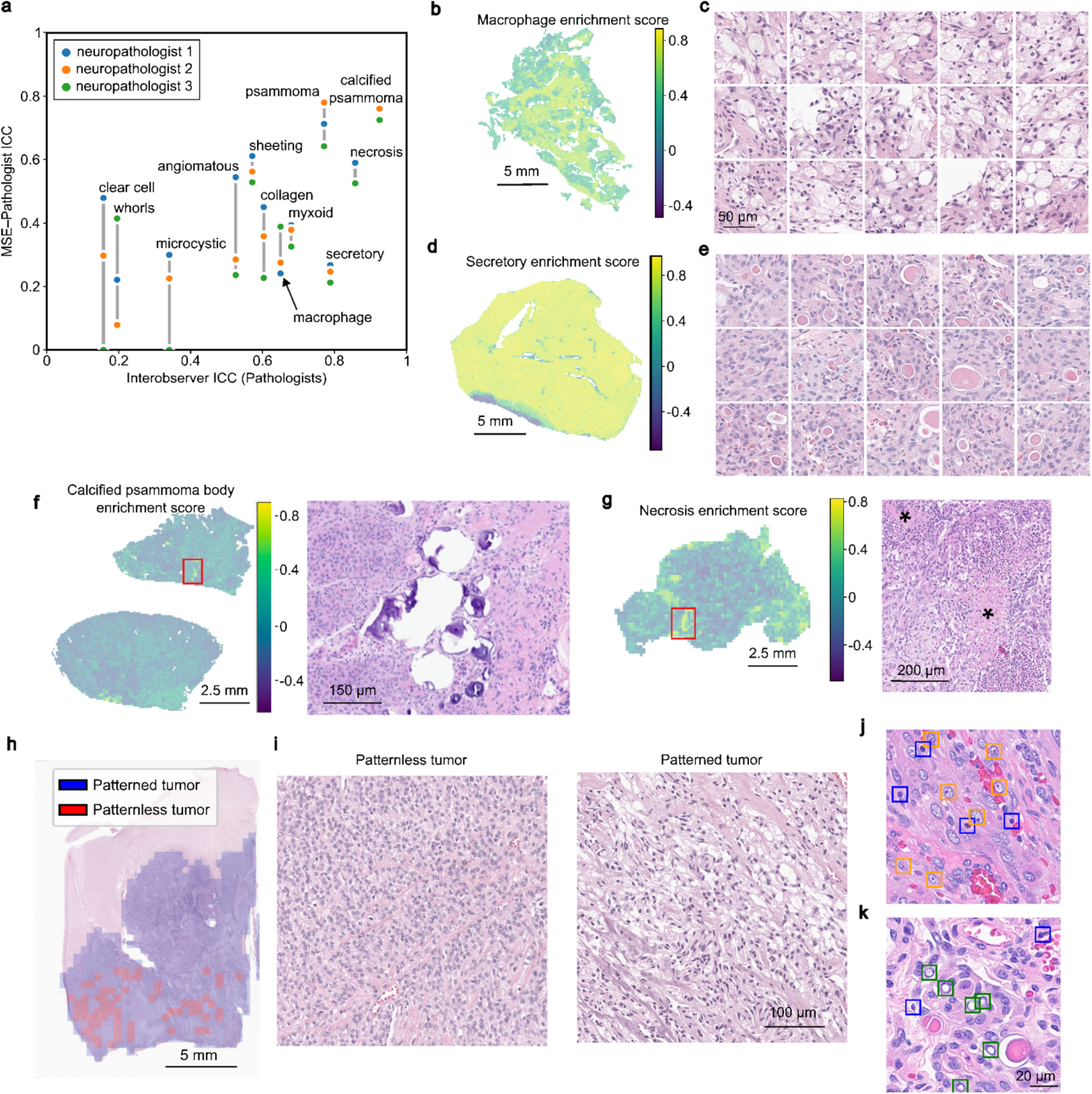
Validating morphology set enrichment. **(a)** We compared MSE scores to ordinal ratings from three board-certified neuropathologists to evaluate pathologist-pathologist and machine-pathologist agreement. **(b)** Heatmap of a macrophage-rich sample highlighting regions with high morphology enrichment (yellow). **(c)** High-power fields with the highest macrophage enrichment scores. **(d)** Heatmap of a secretory-rich sample highlighting regions with high enrichment scores for eosinophilic secretions. **(e)** High power fields with the highest secretory enrichment scores. **(f, g)** Samples with high MSE-pathologist discordance. **(f)** Focal regions with calcified psammoma bodies detected by the MSE algorithm. This sample was scored by pathologists as containing abundant psammoma bodies. **(g)** This sample with focal necrosis was scored as absent necrosis by pathologists. **(h)** Regions with patternless tumor growth, characteristic of sheeting architecture, were quantified by calculating the fraction of low-power fields classified as patternless tumor across the WSI. **(i)** Example of low power fields of classified patternless and patterned tumor growth. **(j)** High power fields showing detections of lymphocytes and macronucleoli, and **(k)** cells with cleared nuclear chromatin (orphan Annie).

Heatmaps visualized enrichment scores and histology features in whole-slide images (**Fig. 3b-i**). A slide containing clusters of macrophages showed hotspots of high macrophage enrichment scores (**Fig. 3b**) associated with clear evidence of clustered macrophages on H&E staining (**Fig. 3c**). Similarly, a slide containing widespread secretory globules exhibited uniformly high secretory enrichment (**Fig. 3d, e**). We further examined slides with low pathologist-machine agreement to understand sources of discrepancies. In a slide that pathologists collectively scored as containing abundant calcified psammoma bodies, only a focal cluster of bodies with coincident high enrichment scores was present. The aggregate enrichment of this slide, however, was low, since this pattern occupied only a small region (**Fig. 3f**). In another slide where pathologists noted the absence of necrosis, enrichment scores clearly indicated necrotic regions that were confirmed by pathologist re-review, highlighting the sensitivity of MSE (**Fig. 3g**). Low specificity of MSE was revealed in some cases, where red blood cells or serum were mistakenly scored as secretory globules (**Fig. S2a-b**). However, in those cases, the enrichment scores were not within the range of correctly identified secretory globules (**Fig. S2c**). A heatmap of HPFs classified as patternless growth (**Fig 3f**, red regions) and representative HPFs with sheeting vs. non-sheeting architecture within the same slide (**Fig. 3i**).

For cell enrichment analysis, embeddings extracted using the pretrained self-supervised model showed clustering of different cell types in a UMAP plot (**Fig. 2e**). Cells classified as lymphocytes, tumor nuclei with macronucleoli, and Orphan Annie nuclei were correctly identified in representative HPFs (**Fig. 3j, k**). These classifications were confirmed by neuropathologist review.

### Unbiased clustering defines morphologic subtypes of meningioma with biologic and clinical correlations

We next performed a cluster analysis using MSE scores (**<u>Table S1</u>**) to examine the morphologic landscape of meningiomas (**Fig. 4**). We identified six salient clusters using objective clustering quality metrics (**Fig. S3a**), defined as sheeting, necrotic, collagenous, lymphocytic, vascular, and classic (**Fig. 4a, b, <u>Table S2</u>**). These clusters do not directly reproduce existing WHO histologic subtypes (**Fig. 4c**) but provide an alternative perspective based on quantitative MSE scores. A UMAP visualization of the heatmap showed that samples group according to the assigned clusters and further revealed additional subclusters (**Fig. 4d**). These clusters can be categorized as poor prognosis clusters (necrotic, sheeting), immunologic clusters (lymphocytic and collagenous with macrophages), a vascular cluster with clear cells, and a cluster with classic meningioma morphological patterns (**Fig. S3c, d, <u>Table S3</u>, <u>Table S4</u>**). These clusters showed significant associations with molecular biomarkers and clinical data (**Fig. S3b, <u>Table S5</u>**).

**Figure 4.**
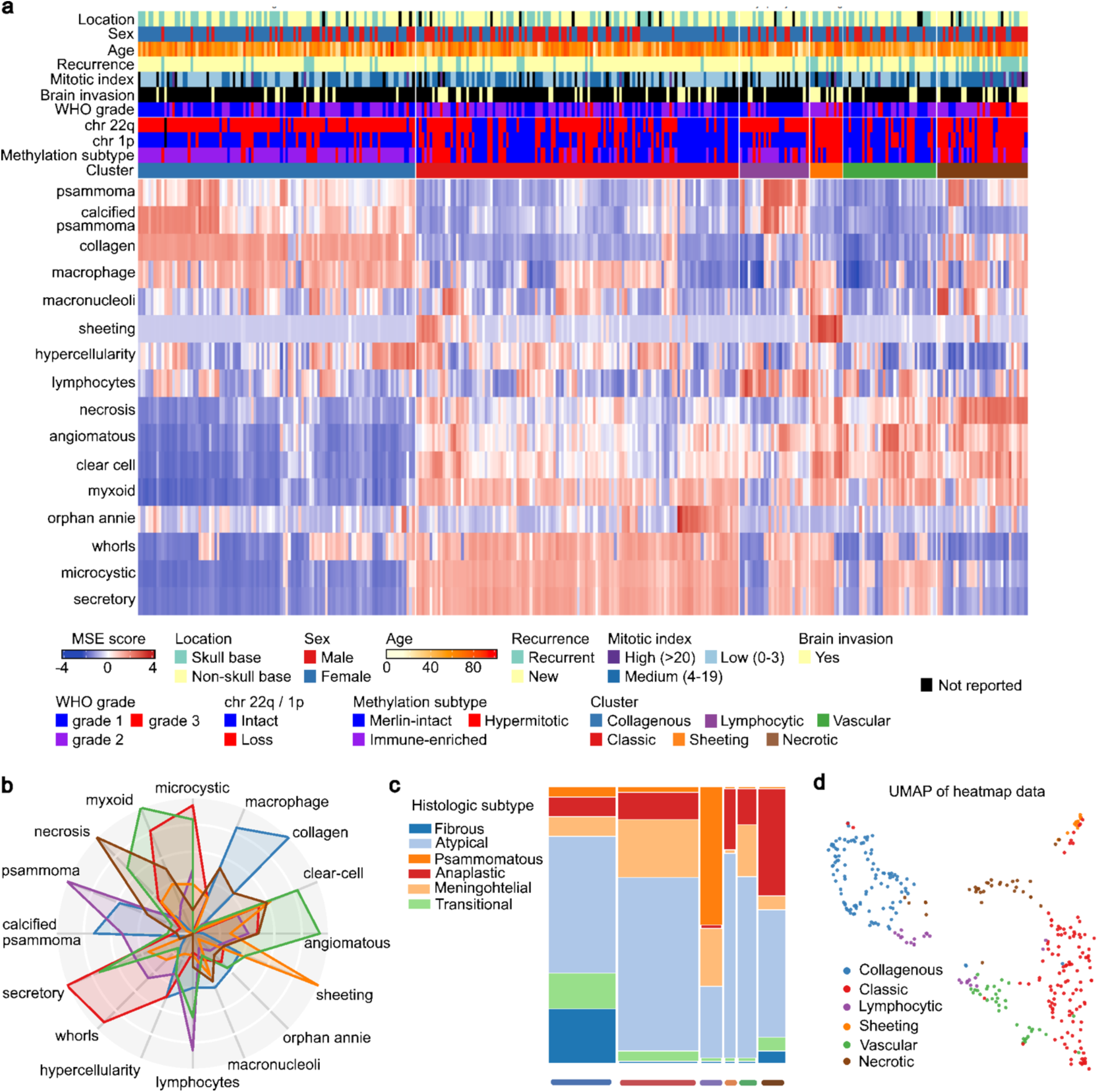
Clustering of morphological set enrichment scores. **(a)** Six clusters emerged labeled as collagenous, sheeting, necrotic, classic, and lymphocytic. Tracks contain clinical and molecular data (**b)** Radar plot of the morphologic patterns associated with each cluster. **(c)** Mosaic plot demonstrating distribution of WHO histological subtypes across the five clusters. **(d)** UMAP of heatmap data with 6 clusters.

The necrotic and sheeting clusters showed shorter progression-free survival compared to other clusters (log-rank test p= 2.34 × 10^-10^) (**Fig. S4**), and showed enrichment for necrosis, macronucleoli, sheeting, and clear cell patterns. Tumors in the necrotic and sheeting clusters predominantly belonged to the hypermitotic methylation subtype (73.3% and 58.1%), with chromosome 1p loss (80.6 % and 86.7%), and 22q loss (80% and 83.9 %). Meningiomas in those clusters had higher recurrence rate (58.7%; vs. 12.5% for other clusters), higher rate of belonging to the high mitotic class (19.6%; vs. 3.1%), higher incidence of brain invasion (43.5%vs. 17.4%), and higher WHO grade (53.1% total WHO grade 3). Many tumors within the sheeting cluster exhibited a weaker signal of classic morphologic patterns (whorls, microcysts, and secretory) while necrotic cluster tumors lacked such patterns entirely. A subset of necrotic tumors showed intense enrichment of macronucleoli in their tumor cells, while such enrichment was rare in other clusters.

The MSE scores also revealed two distinct immunologic microenvironments. The collagenous cluster enriched with collagen, macrophages, and calcified psammoma bodies, while the lymphocytic cluster showed lymphocytic infiltration and was enriched in both psammoma bodies and calcified psammoma bodies. Collagenous and lymphocytic clusters had similarly high rates of being chromosome 1p intact (80.8%, 84.8%), yet also had high rates of 22q loss (92.3%, and 75.8%). These morphologic clusters differed in their proportions of immune-enriched DNA methylation subtype (81% vs 45.5%, respectively). Correlating with standard WHO classification, the collagenous cluster was associated with WHO fibrous subtype (p=4.64 × 10^-4^), while the lymphocytic cluster was associated with psammomatous meningiomas (p=1.6 × 10^-14^).

Clusters with favorable molecular biomarker status included the vascular cluster, enriched with angiomatous, clear cell, and myxoid patterns; this cluster was predominantly merlin-intact methylation subtype (71.4%) with intact chromosomes 1p (65.7%) and 22q (68.6%). The classic cluster, enriched with whorls, secretory, and microcystic patterns, was mostly chromosome 1p intact (65.6%) and predominantly merlin-intact methylation subtype (55.7%). Although the vascular cluster had a higher likelihood of benign molecular biomarkers, it also had shorter progression free survival than the collagenous and immunologic clusters (median rPFS of 927 vs. 2518 days, log-rank test p=0.046). Details of statistical associations of all clusters and clinical and molecular data, using chi-squared tests, are shown in **Fig. S5** and **<u>Table S6</u>**, where Pearson residuals were used to quantify over- and under-representation of cluster-clinical endpoint combinations, and statistical significance was evaluated using Benjamini-Hochberg false discovery rate correction.

### Morphology enrichment predicts DNA methylation subtypes

We incorporated a genomic DNA methylation classifier that consists of three DNA methylation subtypes: merlin-intact; immune-enriched; hypermitotic [16]. Merlin-intact meningiomas have the best outcomes among the three groups and are characterized by intact NF2/Merlin signaling and minimal copy number changes. Immune-enriched meningiomas have intermediate outcomes and are distinguished by immune cell-infiltration, lymphatic vessels, and loss of chromosome 22q. Hypermitotic meningiomas have the poorest outcomes, and exhibit convergent genetic and epigenetic mechanisms that drive aggressive biology, including concurrent losses of chromosomes 1p and 22q [16, 54, 55].

A simple, random forest model based on MSE scores of 16 morphologic patterns was then compared with MIL models for their ability to predict DNA methylation subtypes, finding comparable balanced accuracies (0.784 for MIL and 0.821 for MSE scores) and AUC (0.926 for MIL vs. 0.909 for MSE scores). Differences in AUC performance were not significant (DeLong’s test, BH correction p= 0.8,0.17, 0.8 for merlin-intact, immune-enriched, and hypermitotic, respectively, **<u>Table S7</u>**) (**Fig. 5a, Fig S6.**).

**Figure 5.**
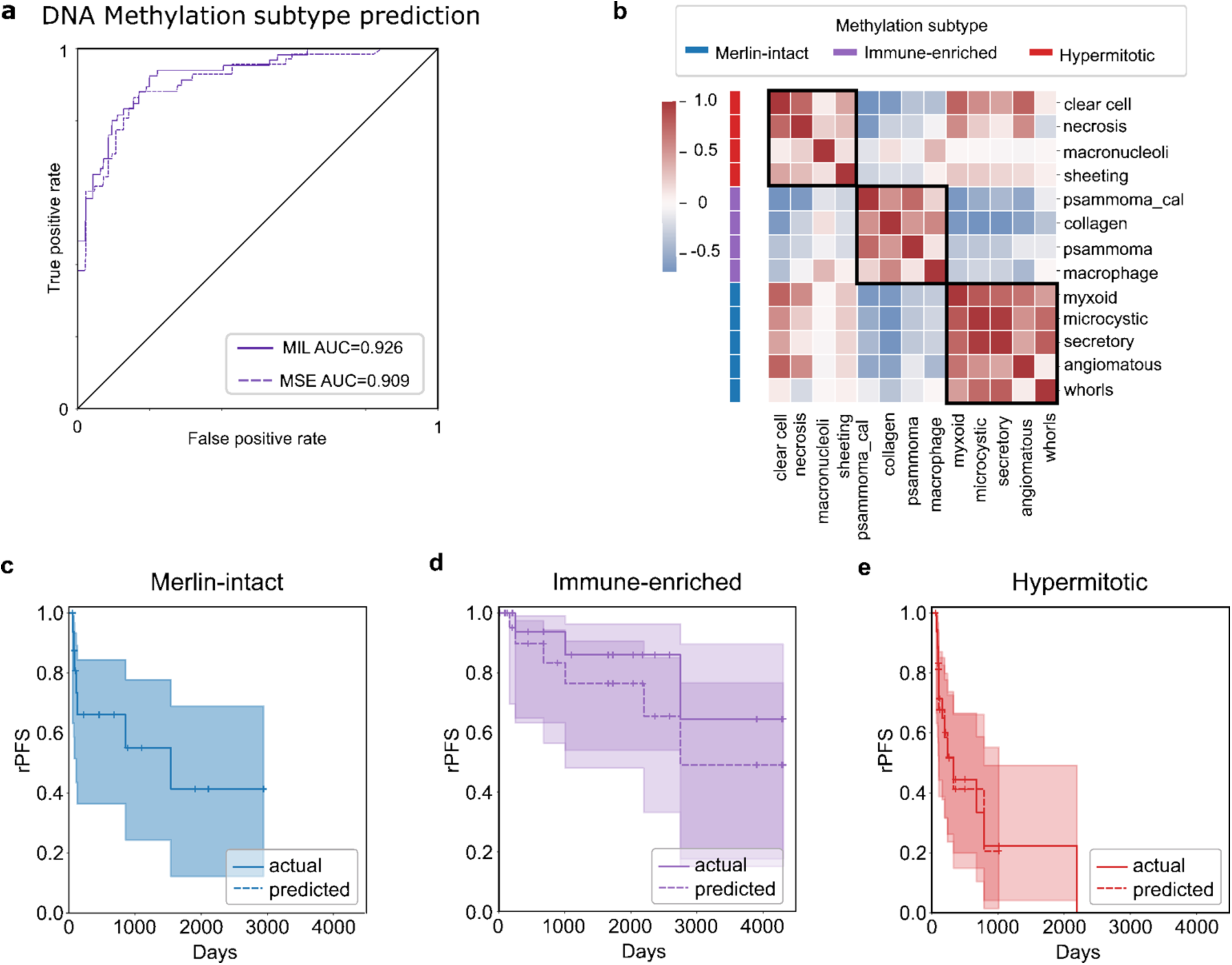
Histology prediction of DNA methylation classification. **(a)** Receiver operating characteristic (ROC) curves for DNA methylation subtype prediction for MSE and MIL models. **(b)** Dominant histology patterns associated with DNA Methylation subtypes. Color indicates inter-pattern correlations. **(c-e)** Kaplan-Meier plots for MSE-predicted vs. actual DNA methylation subtype.

Associations between morphologic patterns and DNA methylation subtypes were assessed using Kruskal-Wallis test with false discovery rate (FDR) correction using the Benjamini-Hochberg method (**Fig. 5b**, **<u>Table S8</u>**). Many morphologic patterns showed a positive association with multiple subtypes. To identify patterns characteristic of DNA methylation subtypes, we assigned each morphologic pattern to a single subtype based on the strongest positive association. Clear cells and macronucleoli were also characteristic patterns of hypermitotic methylation tumors. Collagen, psammoma bodies, calcified psammoma bodies, and macrophages were characteristic patterns of the immune-enriched methylation subtype. The benign merlin-intact methylation subtype tended to have more whorls and microcystic, myxoid, secretory, and angiomatous patterns.

Characteristic patterns of the immune-enriched subtype were conspicuously depleted within other subtypes. Multinomial logistic regression analysis, with significance assessed using FDR-corrected Wald’s tests, revealed that macronucleoli enrichment distinguished hypermitotic from merlin-intact subtypes, while enrichment of clear cell morphology distinguished hypermitotic from immune-enriched subtype, and myxoid distinguished merlin-intact from immune-enriched (**Fig. 5b**, **Fig. S6d-f, <u>Table S9</u>**).

To evaluate the prognostic accuracy of methylation subtype predictions, Kaplan-Meier plots for progression-free survival were compared for actual vs. predicted DNA methylation subtypes. Near perfect alignment in merlin-intact and high overlap in both hypermitotic and immune-enriched were found (**Fig. 5c-e**).

### Morphology enrichment predicts losses of chromosomes 22q and 1p

Transparent models based on MSE scores showed comparable performance to MIL in predicting loss of chromosome 22q and 1p, both in terms of balanced accuracy (22q 0.819 MIL vs. 0.822 MSE; 1p 0.824 MIL vs. 0.83 MSE) and AUC (22q 0.914 MIL vs. 0.898 MSE; 1p 0.922 MIL vs. 0.896 MSE) and (**Fig. 6a, d, Fig. S7**). Differences in AUC performance were not significant between MIL and MSE (DeLong’s test p= 0.67, for 22q and p=0.26 for 1p, **<u>Table S10</u>**). Comparisons of KM plots for predicted and actual losses showed substantial overlap for 1p chromosomal loss, indicating comparable prognostic power between molecular testing and morphology-based predictions (**Fig. 6b, e**).

**Figure 6.**
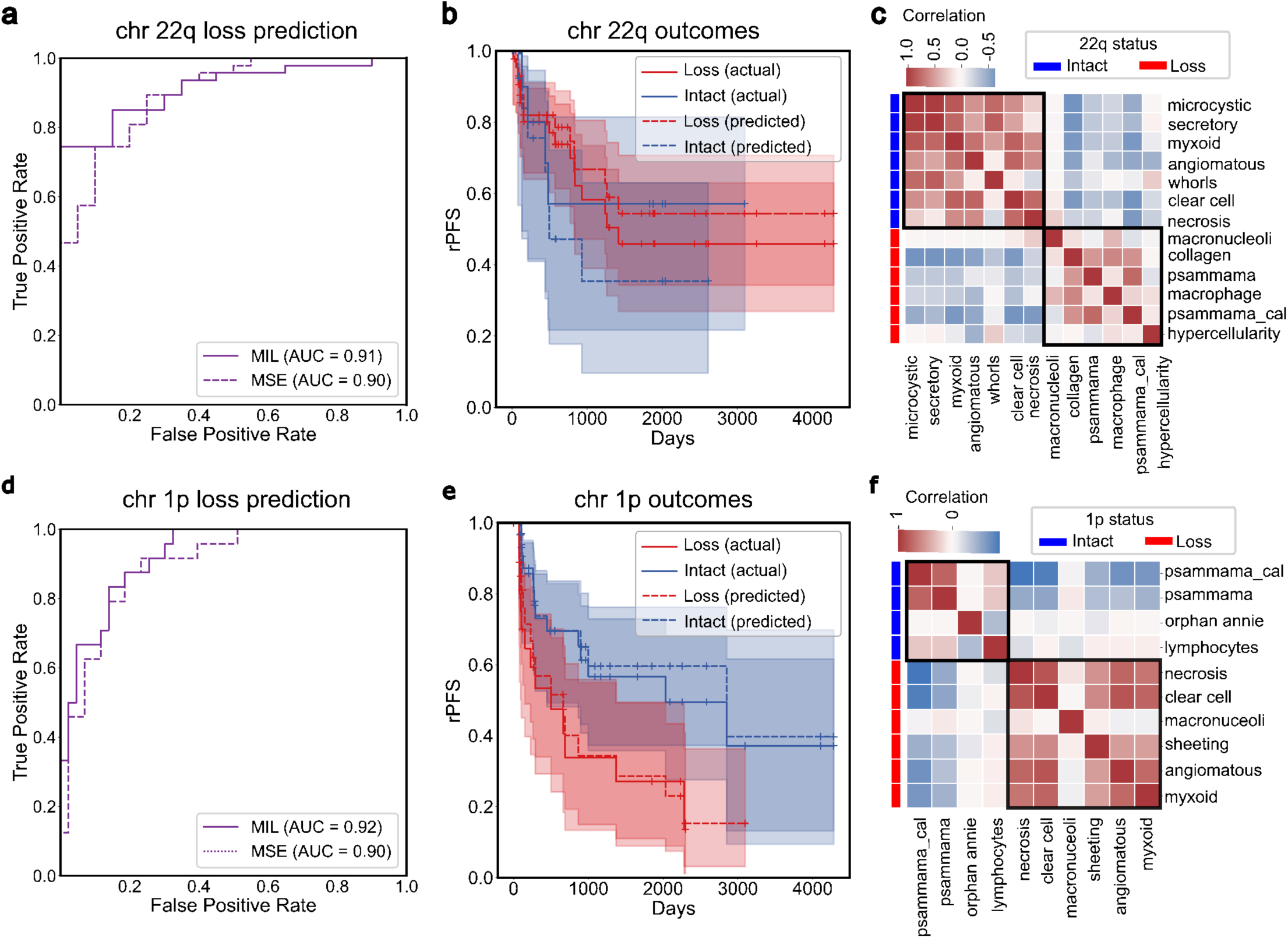
Histology prediction of CNV loss. **(a)** ROC curves for chromosome 22q loss for MSE and MIL models. **(b)** Kaplan-Meier plot for MSE-predicted vs. actual chromosome 22q loss. **(c)** Dominant histology patterns associated with chromosome 22q loss. Color indicates inter-pattern correlations. **(d)** ROC curves for chromosome 1p loss for MSE and MIL models. **(e)** Kaplan-Meier plot for MSE-predicted vs. actual chromosome 1p loss. **(f)** Dominant histology patterns associated with chromosome 1p loss.

Characteristic patterns associated with chromosomal losses, evaluated using Mann U-Whitney tests (**<u>Table S11</u>**), and their pairwise correlations are presented in **Fig. 6c, f**. The characteristic patterns for CNV 22q loss included collagen, macrophages, psammoma bodies, calcified psammoma bodies, hypercellularity, and macronucleoli (**Fig. 6c**). Angiomatous, clear cell, myxoid, necrosis, macronucleoli, and sheeting were characteristic of 1p loss (**Fig. 6f**). Multinomial regression, with significance assessed using FDR-corrected Wald tests, revealed that macronucleoli enrichment is a predictor of chromosome 22q loss (**Fig. S8a, <u>Table S12</u>**). Clear cell and macronucleoli enrichment were associated with 1p loss, while lymphocyte depletion was associated with chromosome 1p intact samples; however, after correction for multiple testing, only clear cell enrichment remained significant (**Fig. S8b, <u>Table S13</u>**).

### MSE improves clinical prognostication

A Random Survival Forest (RSF) was then trained using features from 233 patients in the pattern cohort and validated in 220 patients from the discovery cohort, predicting radiographic progression free (rPFS) survival for primary tumors (**<u>Table S14</u>**, **<u>Table S15</u>**). A baseline clinical model including patient sex and age, WHO grade, mitotic index, and brain invasion was compared with a model based on quantitative MSE scores. For both models, predicted risks were used to stratify into high and low risk groups based on a risk threshold corresponding to the top 20^th^ risk percentile of the pattern cohort, consistent with reported WHO grade proportions (**Fig. S9a**). The MSE model showed better stratification between risk groups (log-rank test p=4.79×10^-5^ vs. 0.143) (**Fig. 7a, b**), with a hazard ratio twice that of the clinical model (HR =2.98 vs. HR =1.48) (**Fig. 7a, b**). By time-dependent Brier score analyses, the MSE model had markedly better performance at 3 years and beyond when compared to the clinical model (MSE-RSF IBS=0.193 vs. clinical-RSF IBS=0.204) (**Fig. 7c**). Combining clinical variables and MSE scores in an RSF model did not improve performance (HR=2.62, IBS=0.192) (**Fig. S9b, c**). Feature importance analyses showed that sex and WHO grade were the most impactful in the clinical model (**Fig. S9d, <u>Table S16</u>**), while lymphocytes, necrosis, and clear cells were the top contributors in the MSE model (**Fig. S9e, <u>Table S17</u>**). Of note, outcomes of subjects from the pattern cohort, who represented the clinical population at Northwestern Memorial Hospital, were generally more favorable than discovery cohort subjects whose tumors had been previously selected for genomic analyses during routine workup (**Fig. S10a-c**).

**Figure 7.**
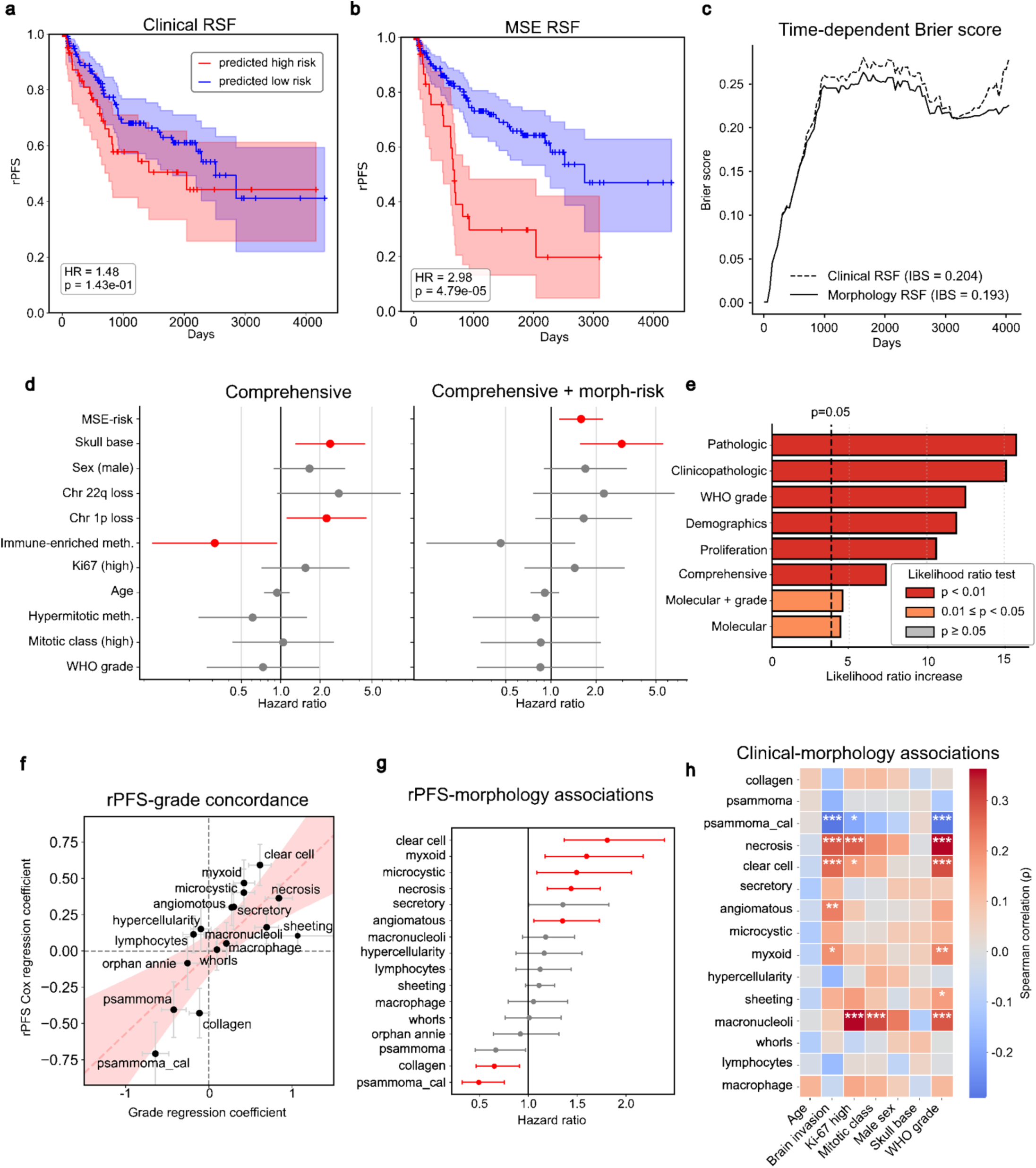
MSE-based outcome stratification. We compared prognostic models based on MSE scores and standard clinical and pathologic variables. **(a)** Kaplan-Meier plot of predicted high- and low-risk patients using clinical variables (WHO grade, age, sex, brain invasion, mitotic class). **(b)** Kaplan-Meier plot of predicted high- and low-risk patients using MSE scores. **(c)** Time-dependent Brier scores comparing clinical- and MSE-based models. **(d)** Multivariable Cox regression of clinical and molecular biomarkers showing MSE-RSF risk values (MSE-risk) provide prognostic value independent of established factors. Red indicates statistical significance after multiple test correction. **(e)** Likelihood ratio increase when combining MSE-risk with other sets of clinical and pathologic variables, demonstrating significant incremental prognostic value over all clinical and molecular variables. **(f)** Association of MSE scores with grade (ordinal regression) and rPFS (Cox regression). Fitted line and confidence intervals shown in red. **(g)** Univariable Cox regressions revealing associations between rPFS and MSE scores. **(h)** Spearman correlations between morphologic patterns and clinical variables (FDR *p<0.05, **p< 0.01, ***p<0.005).

To assess whether the MSE-RSF risk values (MSE-risk) provided prognostic information independent of established clinical and molecular variables, multivariable Cox regressions were performed within the discovery cohort using different combinations of age, clinical, and pathologic variables (**Fig. S11**). MSE-risk was an independent predictor of rPFS in every regression we evaluated. In the comprehensive model containing all variables, both MSE-risk and skull-based location remained independently associated with rPFS (**Fig. 7d**) and significantly improved the likelihood ratio of each regression (**Fig. 7e**). In this study, AI-based mitosis detection was technically difficult, so mitotic index was extracted from pathology reports. However, proliferative markers, including mitotic index and Ki-67, were not prognostically significant apart from the MSE scores. In the comprehensive model, chromosome 1p loss and skull-based tumor location were both associated with higher hazard ratios, indicating shorter progression-free survival, and skull-base tumor location remained significant after adding MSE-risk. The significance of MSE-risk was smallest when combined with molecular variables, further demonstrating the tight relationship with MSE-risk and adverse molecular markers. These results suggest that while MSE scores capture chromosome 1p loss, they do not fully account for the impact of tumor location. Statistical results of all models are shown in **<u>Table S18</u>**.

To identify morphologic patterns that influence rPFS but are under-represented in current WHO grading criteria, the strength of associations between morphologic patterns and WHO grade and with rPFS were compared (**Fig. 7f**). For each morphologic pattern, we fit a univariable logistic regression model for grade prediction (Grade 1 vs. Grades 2,3), and univariable Cox regression model for the rPFS prediction and plotted the corresponding coefficients jointly to analyze their relationships (**<u>Table S19</u>**). Most morphologic patterns closely followed a linear relationship between WHO grade and rPFS. Several morphologic patterns, though, deviated from this trend, suggesting a stronger association with grade than prognosis. Specifically, clear cell showed stronger association with shorter rPFS than with higher grade. In contrast, sheeting showed higher association with WHO grade but weaker association with shorter rPFS. Among indolent morphologic patterns, psammoma bodies aligned closely with the shared grade-survival axis, while calcified psammoma bodies showed a larger deviation, consistent with a stronger protective effect on rPFS than implied by WHO grade. Similarly, collagen showed a modest association with lower grade but a stronger association with better rPFS. Likewise, collagen was associated with better outcomes (p=0.003) (**Fig. S12a, <u>Table S20</u>**), within each grade. KM plots stratified by top 20^th^ percentile of collagen enrichment scores further confirmed that, within each WHO grade, higher MSE collagen scores corresponded with lower risk (**Fig. S12b**).

Using univariable Cox regression, we examined associations between MSE scores and rPFS in the pattern cohort, finding that clear cell, myxoid, microcystic, necrosis and angiomatous (p=5.6×10^-4^, 0.012, 0.036, 0.0012, and 0.037, respectively) were significantly associated with worse outcomes, while collagen and calcified psammoma bodies (p=0.036, 5.9×10^-3^) were associated with better outcomes (**Fig. 7g, <u>Table S21</u>**). These associations trended in the same directions within the discovery cohort (**Fig. S10d, <u>Table S22</u>**).

Associations were then measured between MSE scores in the pattern cohort and clinical variables (WHO grade, age, brain invasion, Ki-67 ≥ 5%, mitotic class (stratified as in WHO grading scheme), male sex, and skull based meningiomas (**Fig. 7h**, **Table 23**). Necrosis, clear cell, and macronucleoli were significantly correlated with high-risk clinical features (WHO grade, mitotic class, and high Ki-67), while calcified psammoma bodies were negatively correlated with these features. Brain invasion correlated with necrosis, clear cell, myxoid, and angiomatous, which may explain why some otherwise “benign” features were associated with poor outcomes in statistical analyses.

Finally, Cox regression with pairwise interaction of MSE scores for rPFS prediction revealed multiple potential interactions among morphologic patterns (**Fig. S12c**), but following multiple testing correction, only the interaction involving necrosis and lymphocytic infiltration remained significant (**Fig. S12d, <u>Table S24</u>**). The prognostic implications of lymphocytic infiltration were strongly modified by the degree of necrosis. As necrosis increased, the hazard associated with lymphocytic infiltration decreased. In tumors with low necrosis, higher lymphocytic infiltration was associated with shortened rPFS. However, in highly necrotic tumors, this association was attenuated and approached zero over time (**Fig. S12e**).

## DISCUSSION

Meningiomas were grossly described in the biomedical literature in 1614 [56], followed by detailed microscopic descriptions in 1864 and 1922 [57, 58]. They are among the most histologically diverse tumors, with 15 variants recognized by the 2021 WHO classification [7]. While histopathology remains the gold standard for prognostic stratification and therapeutic decision-making, grading and subtyping these tumors depends on qualitative and semiquantitative observations that are subject to substantial interobserver variability. Moreover, many of the features and patterns characteristic of meningiomas still have unknown biological significance. This is in part because some morphologic features that might be prognostic, e.g., the number of whorls or Orphan Annie nuclei per mm^2^ in an entire tumor, are impossible to quantify by traditional approaches. Other features that have long been connected to more aggressive behavior, such as macronucleoli, are still subject to interpretive heterogeneity because it remains unclear exactly what proportion of tumor cells have to contain such features to qualify. While screening for specific chromosomal losses and methylation subtypes can provide additional prognostic information, access to these tests remain sporadic. It is also unclear exactly which morphologic patterns are most predictive of adverse molecular biomarkers. As a result, current recommendations for when to consider adjuvant molecular diagnosis in meningiomas are rather complicated [24].

Computational pathology offers a quantitative framework for characterizing the morphologic landscape of meningiomas. This field has grown rapidly, with hundreds of papers describing tools for diagnosis [59–65], prognostication [66–68], prediction of molecular biomarkers [29–33], histological biomarker quantification [69–72], and treatment response prediction [73–76]. However, relatively few studies have focused on meningiomas. Our current work represents a comprehensive investigation that rigorously quantifies the morphologic landscape of meningioma and establishes associations between morphologic patterns, molecular biomarkers, and clinical outcomes. Furthermore, we show that simple interpretable models based on well-established morphologic patterns are highly predictive of underlying molecular alterations and are prognostic of progression-free survival, with accuracy comparable to complex “black-box” models that represent the current dominant paradigm in computational pathology.

We developed MSE, a technique for characterizing morphologic patterns that enables the development of predictive but understandable models. MSE measures the over- or under-representation of morphologic patterns in whole-slide images by statistical analysis of a latent embedding space. This facilitates a comprehensive study of meningiomas while avoiding the extensive work required for developing supervised learning classifiers. MSE scores represent enrichment or depletion of morphologic patterns relative to a large baseline, which is helpful in studying meningiomas and other diseases where multiple morphologic patterns can be present to different degrees in a given tumor. MSE scores are summarized at the patient level, but spatially resolved scores are also generated to enable visualization. When validated against neuropathologist annotations, MSE scores showed comparable concordance to that observed among pathologists. Analysis of discordant cases found that, in some cases, MSE detected the presence of morphologic patterns that were marked by neuropathologists as absent, e.g. necrosis, suggesting that MSE provides high sensitivity. We also found that specificity can be a weakness of MSE, noting that red blood cells were sometimes mischaracterized as secretory globules, and that MSE was unable to discriminate patterned vs. patternless tumor growth. Future work will address this by exploring how to incorporate negative examples of morphologic patterns in the MSE framework. Still, some patterns exhibiting low machine-pathologist agreement, including clear cell and macrophages, were subsequently shown to be highly predictive of molecular biomarkers. This suggests that low agreement does not necessarily imply machine errors, but rather that MSE can capture meaningful signals that are not reliably scored by pathologists.

A major strength of MSE is its interpretability and transparency. Because the method quantifies interpretable morphologic patterns at scale, it enables statistical identification of prognostically relevant patterns that have been previously overlooked in WHO grading. Although many morphologic patterns have similar degrees of association with WHO grading and rPFS, their relative contributions to prognosis differ. Specifically, clear cell showed a stronger association with poor outcomes than is currently reflected in WHO grading. This signal is not a recapitulation of the clear cell histological subtype, which requires more than 50% clear cell morphology on the slide. Our discovery cohort contained only 2 tumors formally diagnosed as clear cell meningiomas, yet 25 patients exhibited elevated clear cell MSE scores. Similarly, collagen deposition remained a protective pattern even within higher grade tumors, suggesting that prognostic significance is independent of tumor grade. This signal is also not attributable to the fibrous meningioma subtype, since 14 patients were diagnosed with fibrous meningiomas, while 128 patients exhibited elevated collagen MSE scores. Calcified psammoma bodies showed a stronger protective association than psammoma bodies in general, suggesting higher chronicity indicates a more favorable prognosis. In contrast, sheeting carried more weight in WHO grading but showed less association with rPFS. Collectively, these results demonstrate that multiple morphologic patterns influence prognosis but are difficult to quantify reproducibly by manual histological review and, therefore, may not be amenable to current grading schemes. Although clinical deployment of AI models remains challenging due to the requirements for accuracy and generalizability across hospitals, the interpretability of MSE reveals key morphologic patterns that could be incorporated into improved WHO grading criteria.

We present for the first time an unbiased clustering of meningiomas into six groups based on objective and quantitative morphologic patterns. This provides a new perspective on how meningiomas self-aggregate in terms of morphologic patterns, in contrast to established histologic subtypes. The sheeting and necrotic clusters have poor prognosis and associations with higher histologic grade, brain invasion, and mitotic index, and predict hypermitotic DNA methylation subtype and concurrent losses of chromosomes 1p and 22q. While sheeting as an individual morphologic pattern was more strongly associated with WHO grading than with rPFS, the sheeting cluster demonstrated significantly worse rPFS. This suggests that the prognostic impact of sheeting arises from co-occurrence with additional adverse morphologic features, rather from sheeting alone. Supporting this, we identified a subcluster within the classic cluster characterized by lower sheeting MSE scores (relative to the sheeting cluster) and a combination of several classic morphologic patterns, which had more favorable outcomes compared to the sheeting cluster. This may reflect differences between contiguous vs. patchy sheeting across the WSI.

A cluster characterized by dense collagen, macrophage infiltration, and calcified psammoma bodies was predominantly immune-enriched and exhibits chromosome 22q loss with relative preservation of 1p, indicating a less proliferative yet chronically inflamed tumor microenvironment. Indeed, the immune-enriched methylation subclass was prominent within the collagenous cluster, suggesting that, as is the case elsewhere in the body, chronic inflammation may lead to macrophage-driven fibrosis in the stroma and whorls of meningiomas, in turn accelerating the development of calcified psammoma bodies from those whorls.

The lymphocytic cluster, marked with lymphocyte infiltration and associated with psammoma bodies and calcified psammoma bodies, showed the most favorable rPFS, although differences relative to collagenous and classic clusters did not reach statistical significance, likely due to a limited number of events. This cluster also showed associations with intact chromosome 1p and chromosome 22 q loss. While many lymphocytic cluster tumors are classified as immune-enriched methylation subtype, unlike the collagenous cluster, the cluster is not predominantly immune-enriched. This may reflect the fact that immune-enriched methylation subtype captures a coordinated tumor-level program that includes immune cell infiltration, reduced tumor purity, tumor cell HLA class II expression, lymphatic vessel formation and extracellular matrix remodeling, rather than lymphocyte abundance alone [16]. Accordingly, tumors in the lymphocytic cluster can therefore show consistent lymphocyte presence without engaging in the full epigenetic program required to shift global methylation subtype. Furthermore, the lymphocytic cluster is not a single biological state, it consists of at least 2 subclusters as evident in the UMAP plot (**Fig. 5d**), which likely dilutes its correspondence with the immune-enriched methylation subtype. These findings are broadly consistent with Maas et al. [28] that showed higher immune content in benign meningiomas, reflected in our results as macrophage-rich HPFs with 1p intact and 22q loss tumors. Furthermore, Maas et al. showed that the immune-enriched methylation subtype as defined by Choudhury et al.[16] is more common in less aggressive cases. However, our results suggest that immune infiltration alone does not fully explain tumor aggressiveness, and that morphologic patterns like necrosis, clear cells, and calcified psammoma bodies can attenuate the prognostic influence of tumor-associated macrophages. Although methylation subtyping is widely used in neuro-oncology, the imposition of discrete classifications presents a problem where risk related patterns exist on a continuum. The ability to capture these patterns in a quantitative manner presents a new opportunity for understanding the tumor evolution, chronicity, immune dynamics, stromal remodeling, and other tissue remodeling changes.

Overall, MSE scores highlighted clear cell as a morphologic pattern consistently associated with adverse molecular biomarkers or worse outcomes, while calcified psammoma bodies and collagen were protective patterns. These morphologic patterns may represent prognostically meaningful features that are currently underweighted in WHO grading.

Using MSE scores, we trained a survival model to predict rPFS. This model achieved substantial improvement in predictive performance compared to a baseline model trained on routine clinical variables (WHO grade, age, sex, mitotic index). MSE-risk was independent of all other variables including demographics, proliferation, pathology, and molecular biomarker status. Although mitotic index is one of the most important histologic variables in current WHO grading, it was not prognostically independent of MSE-risk, suggesting that it did not add further value to the MSE-risk score. Nevertheless, one future direction will be to incorporate AI-based mitotic counts into the algorithm, rather than relying on indices extracted from pathology reports. Likewise, future iterations of this system will explore whether AI-based radiologic imaging analyses further refine the prognostic accuracy of our MSE-risk.

In summary, the MSE approach can be extended beyond meningiomas as a tool to predict survival and molecular biomarkers while generating biologically grounded hypotheses by quantifying the morphologic landscape of meningiomas. The AI models presented in this study underscore the interplay between morphology, genomic alterations, and epigenetic states, suggesting that rigorous quantification of morphologic patterns can enhance our understanding of molecular pathways and clinical behavior. Furthermore, MSE is interpretable, which presents opportunities to identify patterns that can improve grading criteria, providing a clearer path to clinical translation.

## METHODS

### Clinical datasets

We divided our data into a pattern cohort and a discovery cohort. Both include WSIs, clinical data including patient age at time of resection, progression free survival assessed by radiologic imaging (rPFS), WHO grade, mitotic index, and partial information on brain invasion, skull base location, and WHO histologic subtype (**Table S2**). In our analyses mitotic index was stratified to low (< 4 / HPF), medium (4-19 /10 HPF) and high (≥ 20 / 10 HPF). The discovery cohort additionally included DNA methylation subtypes and copy number variation calls for chromosomes 1p and 22q. The pattern cohort was used to develop feature extraction methods, for annotation of morphologic patterns, patternless tumor, and nuclei, and for training of models predictive of clinical outcomes and molecular biomarkers. The discovery cohort was reserved for evaluating predictive models, for measuring the prognostic independence of variables, and for clustering morphologic patterns (**Table S2**).

### Acquisition and analysis of DNA methylation data

DNA was extracted from FFPE or frozen tissue specimens where DNA methylation data was obtained on the Illumina platform using the Infinium MethylationEPIC (EPIC v1) or Infinium MethylationEPIC v2.0(EPIC bv2) array. IDAT format files were preprocessed and normalized using the SeSAMe R package (Bioconductor v3.18). A previously reported Support Vector Machine (SVM) classifier with 3 methylation subtypes (merlin-intact, immune-enriched, hypermitotic) was used to stratify patients. Genomic CNV profiles were inferred from methylation profiles by performing copy number segmentation, a function available in the SeSAMe package. Thresholds of inferred segments with mean log intensity less than −0.15 were defined as CNV loss, and segments with mean log intensity between −0.15 and 0.15 were considered CNV neutral (intact).

### Morphologic patterns and patternless tumor HPFs feature extraction

Extracting features from image patches is a preprocessing step required in both the MIL and MSE models. Features were extracted using the UNI2-h foundation model [77] that was trained on 200× magnification image patches from more than 350 thousand diverse WSIs across multiple diseases including neoplastic, infectious, and inflammatory conditions. For MIL training all models were trained with HPFs extracted at 200× magnification with a size of 112 x 112 mm (224×224 pixels). For classification of patternless tumor associated with sheeting 50× magnification 448 x 448 mm fields were embedded using UNI2-h.

### Multiple instance learning models

Multiple instance learning (MIL) models were developed as a baseline to compare against [78]. MIL models make patient-level predictions from collections of HPFs and assign an attention score to each HPF indicating its contribution to the prediction. Feature embeddings were processed independently through a shared backbone network, weighted via attention, and aggregated into a single bag-level representation before passing through a shared classifier head (**Fig. S13**). MIL models were trained using and Adam optimizer with a learning rate of 1×10^-5^, optimized using a hinge-based loss function.

### Annotation of morphological, cellular, and sheeting patterns

To build a reference library for the MSE algorithm, examples of the morphologies of interest were annotated and reviewed by a board-certified neuropathologist before calculating the enrichment scores. Annotation was performed in the pattern cohort using the Digital Slide Archive platform, a web-based platform for management and annotation of digital pathology imaging data [79, 80].

### MSE models for morphological patterns

Morphology set enrichment was inspired by the Gene Set Enrichment Analysis (GSEA) used in bioinformatics analysis [81]. We start by defining the tile-level enrichment score for a single pattern and then extend this to patient level scoring below. The matrix *Q* contains the *f*-dimensional embeddings of *n* HPFs, *q_i_* for *i*=1,…,*n*, from a single slide. The matrix *P* contains the embeddings of annotated HPFs, *p_i_* for *i*=1,…,*m*, for the *m* annotated examples of one pattern. The matrix *R* contains *m* embeddings sampled from a larger pool of tiles drawn randomly from all pattern cohort slides. For each morphology pattern, we pre-sampled 500 random sets, that together define a fixed collection used to calculate similarity statistics under a random sampling condition. These random sets were held constant across all enrichment calculations to ensure consistency across tiles, slides, and patients. Given a query HPF, enrichment scores were computed by comparing its embedding to the pattern HPF examples against each of the random sets in *R*. The equations define the enrichment statistic for a single random draw. Multiple random draws are then used to stabilize the enrichment estimate, and the resulting scores were aggregated to obtain a robust tile-level enrichment score.

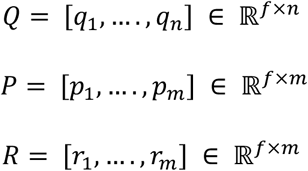

Firstly, cosine similarity is computed between the query slide *Q* and examples *P* from one pattern

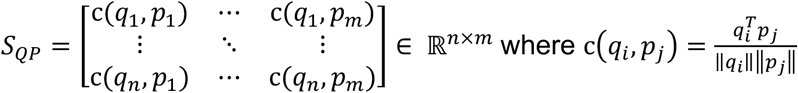

The similarity *S_RP_* between the random sample *R* and the pattern examples is also calculated. The sizes of *P* and *R* are matched at *m* so the concatenated similarity matrices *X* defines a balanced ranked space with equal numbers of hit and miss entries. This matrix is associated with label matrix *L* containing an indicator for *S_QP_* values

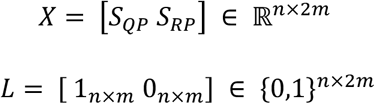

Similarities were ranked for each HPF *i* and the pattern examples *P* along the rows *X_iP_* of *X*. Let *Π*_*i*_ = argsort (*X*_*iP*_) be the descending sort order of *X*_*i*_, and apply this sorting to the labels 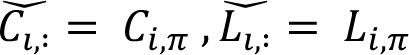. Similar to Gene Set Enrichment Analysis we calculate weighted hit-miss increments at each rank position *k*, weights denoted as 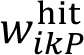 and 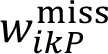 from which normalized hit increments 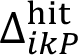 and miss decrements 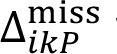 were computed.

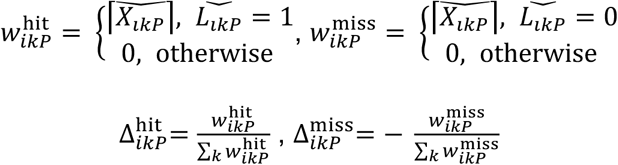

Finally, we calculated the running sum for each HPF, where *t* indexes the cumulative sum over ranked similarities 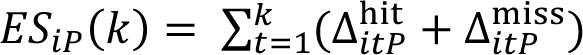. and the total enrichment score for each tile is given by the maximum deviation of this running sum across all *k*.

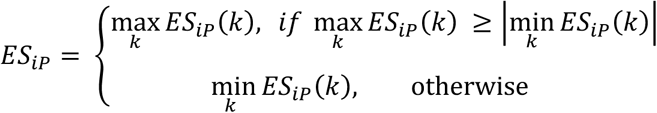

For each HPF, enrichment scores were calculated across the random sets in *R*, and these are aggregated via an average to produce a final stabilized enrichment score per HPF. To identify an optimal method for aggregating morphology enrichment scores across image patches at the patient level, we compared multiple aggregation approaches against a pathologist’s scoring. A pathologist provided ordinal scores (0-3) for each morphology in the pattern library across 18 patients. For each aggregation approach we computed Spearman’s correlation between the aggregated enrichment score and the pathologist’s score (**Fig. S1a**). We evaluated multiple aggregation strategies including: (a) the median of slide-level medians of tile scores(median_medianS),(b) the median of slide-level 75th percentiles of tile scores(median_75S), (c) the median of slide-level medians of tile scores (max_medianS) (d) the maximum of slide-level 75th percentiles of tile scores(max_75S), (e) the overall median of all tiles pooled across slides(median_tiles), (f) the 90th percentile if tile scores pooled across slides(percentile90_tiles). The overall median of all tiles showed the highest correlation and was used for patient-level aggregation.

### Nuclear feature extraction

We used the CellVit++ Python package to detect nuclei from hematoxylin and eosin-stained WSIs [82, 83]. Following nuclear detection, 32 x 32-pixel images capturing 8 x 8 mm regions at 400× magnification were cropped around the centroids of segmented nuclei from 5000 randomly selected cells per slide, resulting in a total of 4,691,209 images. A self-supervised Distillation with No Labels (DINO) model [84] was trained on images using https://github.com/computationalpathologygroup/dino.git. A Vision Transformer (ViT-Tiny) backbone was used, producing 192-dimensional patch embeddings with an input resolution of 32 pixels and token-size of 8. During training, a DINO projection head with 16,384-dimensional output was applied for self-distillation between student and teacher networks. This projection head was used only during training and discarded during later use. The teacher network was updated using an exponential moving average with a base momentum of 0.999. The model was trained on for 50 epochs using AdamW optimizer with a learning rate of 3-5, weight decay of 1×10^-5^ and gradient clipping to a maximum value of 2.0. A cosine learning-rate schedule with linear warm-up over the first five epochs was used, with learning rate decayed to a minimum of 3×10^-6^. Weight decay was scheduled using cosine schedule, increasing from 0.02 to 0.2 over training. The final projection layer was frozen for the first training epoch to improve optimization stability. Models were trained with a batch size of 64 per GPU over 10 L40S NVIDIA GPUs. Multi-crop data augmentation was used during training, consisting of two global views sampled with a scaled range of 0.25-0.5 and eight local sampled with a scale range of 0.25-0.5. The teacher temperature was fixed at 0.04 through training, with no warm-up period. The trained ViT-Tiny was then used to extract feature embeddings for annotated nuclei of the pattern discovery cohort used in classifier development, and for each nucleus within the discovery cohort.

### Cell classifier and hypercellularity count

We trained a cell nuclei classifier to distinguish between lymphocytes, orphan Annie tumor nuclei, tumor nuclei exhibiting macronucleoli, and other tumor nuclei not exhibiting these features. The labeled data was reviewed by a board-certified neuropathologist before training the supervised classifier. Image patches of 437 for lymphocytes, 1970 macronucleoli tumor nuclei, 2062 orphan Annie tumor nuclei, and 2222 other tumor nuclei were annotated and reviewed by a neuropathologist. Feature embeddings(192-dimensional) were used as inputs to a multilayer perceptron (MLP) classifier. The network consisted of 2 hidden fully connected layers with 32 units each. ReLU activations were applied after first and second layers, and dropout (p=0.3) was applied between second and the output layer for regularization. The model output was transformed to class probabilities using a SoftMax function. The labeled data was split into training (80%) and held-out test (20%) sets. Training was performed using an Adam optimizer (learning rate=1×10^-5^) for 30 epochs. A weighted cross-entropy loss function was used to mitigate class imbalance, and class weights were computed using a balanced weighting scheme based on label frequency. Model performance was assessed on the held-out test set. For cell enrichment analysis, all detected cells were inferred using the trained classifier, and cell labels were assigned only when SoftMax scores exceeded the predefined threshold of 0.9 for a single class. For further qualitative validation, a neuropathologist reviewed classified cells. Hypercellularity was defined as the total number of detected nuclei divided by the total analyzed tissue area across all slides for a given patient.

### Sheeting classifier

Three neuropathologists labeled a dataset of 201 patternless HPFs and 854 patterned HPFs from the pattern cohort for a supervised classifier. We trained a supervised classifier of patterned and patternless tumor to detect sheeting growth at 50× magnification. Image embeddings extracted from 448 x 448 mm fields using UNI-2h with 1536 dimension were used as inputs to a MLP classifier. The network consisted of 2 hidden fully connected layers with 64 units each. ReLU activations were applied after first and second hidden layers, and dropout (p=0.5) was applied between second and the output layer for regularization. The model output was transformed to class probabilities using a SoftMax function. The labeled data was split into training (80%) and held-out test (20%) sets. Since the sheeting architecture is rarely available across the meningioma dataset, we used a focal loss with class-specific weighting α= [1, 2], and γ=3. The model was trained using an Adam optimizer with a learning rate of 5×10^-6^ for 70 epochs. We inferred patternless HPFs from the discovery cohort. For validation, neuropathologists assigned an ordinal value (0-3) for a set of 44 slides which we compared with our patient-level sheeting enrichment values.

### Cell and sheeting enrichment analysis

Both the cell morphology and the sheeting enrichment analyses were classifier-based. Following nuclear classification, we computed a smoothed enrichment ratio comparing the observed fraction of the cell type within a patient to the expected fraction in the full cohort. For each patient p and cell type *c*, *k_pc_* is the number of cells of type *c* in patient *p*. And *n_p_* is the total number of cells in patient *p*, *K_c_* is the total number of cells of type *c* across the cohort, and *N* is total number of cells in the cohort. A pseudo-count parameter *α*=0.5 was used to stabilize estimates for rare cell types.

The smoothed observed and expected fractions are defined as

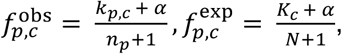

and the smoothed enrichment ratio for each cell is defined as:

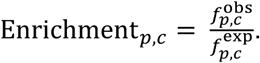

Similar equations are used for calculating enrichment scores of sheeting.

### Neuropathologist scoring analysis

For validation, we selected 3 slides for each morphologic pattern with different DNA methylation subtype and had high MSE scores for that morphology. We additionally selected another slide per morphological pattern with a low MSE score, resulting in a total of 44 validation slides. Neuropathologists were blinded to both the methylation subtype and the corresponding MSE pattern and sheeting scores. They independently assigned an ordinal rating (0-3) for each pattern, which were used to assess agreement with MSE scores. Agreement for each slide-morphology pair was quantified using the intraclass correlation coefficient (ICC), calculated with a two-way mixed-effects model with absolute agreement ICC(3,1) [85] using the *pingouin* Python package. Before analysis, all MSE scores were standardized within each pattern using the *scikit-learn* Python package. We first calculated inter-pathologist ICC per morphology. We then calculated agreement between MSE scores and pathologist scores by computing ICC between standardized MSE scores and each pathologist’s ordinal ratings. For each morphology, the range of MSE scores-pathologist ICC values were compared with the corresponding interobserver ICC among neuropathologists.

### Predictive survival models

We trained a random survival forest (RSF) model to predict progression-free survival using clinical covariates (age at diagnosis, sex, brain invasion status, mitotic class, and WHO grade). Model implementation used Python *scikit-survival* package. For model development, the pattern cohort was randomly split into training (80%) and validation (20%), and the discovery cohort was used for testing. Hyperparameter optimization was performed using randomized search (50 sampled configurations). For each sampled configuration, models were trained with 3,000 trees, bootstrap sampling, and unrestricted tree depth. Parameter combinations in which the minimum split size was less than or equal to the minimum leaf size were excluded to ensure valid tree structures. For Clinical-RSF, the search space included minimum number of samples required at a leaf node (3-20), minimum number of samples required to split and internal node (6-40), and maximum features (square root of the total number of variables, 40% and up to all variables). For the MSE-RSF and the Clinical and MSE-RSF combined we used a restricted search space to larger node sizes to reduce overfitting in higher-dimensional feature spaces. The search space included a minimum number of samples required at a leaf node (8-20), minimum number of samples required to split and internal node (16-40), and maximum features (square root of the total number of variables, 40% and up to all variables). Model performance was evaluated on the validation dataset using Harrell’s concordance index (c-index), computed from predicted risk scores. The best model with the selected hyperparameters was refit on the full pattern cohort to calculate the risk scores: Clinical-risk, MSE-risk, and Clinical-MSE-risk combined.

Risk stratification was performed by defining a high-risk group as patients whose Clinical- or MSE or Clinical-MSE-risk fell within the top 80^th^ percentile of the corresponding distribution in the pattern cohort. This threshold was subsequently applied to the discovery cohort to assign patients to high- and low-risk groups. Kaplan-Meier survival curves were generated to visualize differences between risk groups. Kaplan-Meier plots were then used to assess differences in rPFS between the two groups, and log-rank test (Python *lifelines* package) to calculate the significance. To quantify the effect size associated with risk group membership, a univariable Cox proportional hazards model was fitted with the risk group (high vs. low) as the only covariate. Hazard ratios (HRs) with corresponding 95% confidence intervals were reported.

Model calibration and overall predictive accuracy were assessed using time-dependent Brier score and integrated Brier score (IBS). Survival probability estimates from the final RSF models were evaluated on the discovery cohort over a predefined range of follow-up times, with the pattern cohort used as the reference for censoring adjustment. Integrated Brier scores were computed by averaging the time-dependent Brier score over the evaluation time interval. Brier score curves and IBS values were used to compare predictive performance of Clinical-RSF, MSE-RSF, and Clinical-MSE-RSF models.

To assess the relative contribution of individual features to the final RSF models, permutation-based feature importance was calculated for each RSF (Python *scikit-survival* package). Feature values were randomly permuted 50 times, and the resulting decrease in model performance was measured. Model performance was evaluated using the c-index. Importance values were summarized as the mean decrease in c-index with corresponding standard deviation reflecting variability across permutations. Features were ranked according to their mean permutation importance, and error bars represented standard deviations.

### Independent prognostic performance of MSE-risk

Cox proportional hazards (Python *lifelines* package) was used to evaluate associations between covariates and rPFS. For each covariate set, two multivariable models were fitted: a base model containing the clinical, histological or molecular variables, and an extended model that additionally included a continuous z-scored MSE-risk. Clinical, demographic, pathologic and molecular variable were harmonized prior modeling. Age was scaled per 10-year increase, sex was coded as male vs. non-make, and race was dichotomized as White vs. non-White. Methylation subtype was treated as a categorical variable with merlin-intact as the reference group. WHO tumor grade was treated as an ordinal variable (grades 1-3). Mitotic activity was encoded as an ordered categorical variable (low, medium, high), and skull location was included as a categorical covariate. Models were fit using the same patient subset in the discovery to ensure comparability. Model coefficients were summarized as hazard ratios with corresponding 95% confidence intervals and forest plots were generated to visualize effect estimates for base and extended models.

### Univariable Cox regression and WHO grade concordance

We ran a univariable Cox proportional hazards regression on both the pattern and discovery cohort, with MSE scores as continuous variables. We further extended this analysis to compare the relative weighting of morphologic patterns in WHO grading vs. rPFS. For the discovery cohort we performed a univariable Cox proportional hazard regression for rPFS and a univariable logistic regression coefficient for WHO grade (grade 1 vs grade 2,3) for each morphologic pattern. Regression coefficients from the grade and survival models were compared, and a linear regression was fitted across coefficients to summarize concordance between WHO grade and survival associations.

### Clinical-morphology associations

We calculated Spearman rank correlation between MSE scores and clinical or pathologic variables. Ordinal variables (WHO grade and mitotic class) were encoded as ordered numerical variables. Correlations were computed using pairwise observations. P-values were adjusted for multiple testing correction using Benjamini-Hochberg false discovery rate method. Results were visualized as a correlation heatmap with Spearman correlation coefficients and significance indicated after FDR correction.

### Clustering analysis

We performed hierarchical agglomerative clustering of standardized MSE scores using ward linkage and Euclidean distance, implemented with the R *stats* package. Clustering was applied to both samples and MSE scores. Patient-level clusters were defined by cutting the hierarchal dendrogram to yield six clusters, which were subsequently labeled based on characteristic morphologic patterns. The number of clusters was selected by jointly considering clustering quality metrics. While silhouette score peaked at 5 clusters, the 6-cluster solution retained a relatively high silhouette score while showing continuous improvement in the Davies-Bouldin index. 6 clusters therefore represent the minimum number of clusters that balances cluster separation and compactness across both metrics. For visualization purposes only, standardized feature values were transformed using the inverse hyperbolic sine (arcsinh) transformation prior to heatmap visualization to improve dynamic range, while clustering was performed on the original standardized MSE scores. Heatmaps and annotations were generated using R *ComplexHeatmap* package. A centroid was calculated for each cluster by taking the mean of all morphology enrichment scores across the patients belonging to that cluster. Each centroid defines the representative morphology profile of that cluster and is used to evaluate the characteristic patterns of each cluster. Morphologies with scores exceeding ±1 standard deviation from the mean were classified as over- or underrepresented

We generated Kaplan-Meier plots for each cluster and performed pairwise log rank tests rPFS to assess statistical difference between clusters. To identify clinical, histologic, molecular biomarkers associated with individual clusters, categorical variables were compared across clusters using Fisher’s exact test for binary variables and Pearson’s chi-square test for multiclass variables. Continuous variables(age) were compared across clusters using the Kruskal-Wallis test. To identify which specific cluster-clinical end point combinations contributed to these effects, we computed standardized Pearson residuals for each combination and applied multiple test-correction using the Benjamini-Hochberg method. Significant positive residuals indicate overrepresentation, while negative residuals indicate underrepresentation.

### Methylation subtype classifier

A supervised multi-class Random Forest classifier was trained using MSE scores (patterns, cells, and sheeting). Patients in the discovery cohort (334) stratified by DNA methylation subtype and randomly split into training (80%) and held-out test (20%) sets. The same split was used in training the corresponding MIL model. Model hyperparameters were optimized using stratified five-fold cross-validation on the training samples, with performance assessed using weighted one-vs-one under the receiver operating characteristic curve ROC AUC. Hyperparameters explored included the number of trees (50-800), maximum tree depth (10-30), number of features considered at each split (50% or all available features), minimum samples required for split and leaf nodes, and bootstrap sampling. A total of 150 random hyperparameter combinations were evaluated. Class imbalance was addressed using balanced class weights during model training. The best-performing model was selected based on cross-validated ROC-AUC and subsequently evaluated on the held-out test set. The model performance was evaluated on the test set using class-specific one-vs-rest ROC AUC values, macro-averaged AUC, confusion matrices, and balanced accuracy. All analyses were done in Python *scikit-learn* package. Overall differences in MSE scores across methylation subtypes were assessed using Kruskal-Wallis tests with pairwise comparisons performed using Benjamini-Hochberg adjusted Dunn’s test. Delong’s test was used to compare ROC-AUC curves from MSE-scores vs. the MIL results[86]. For visualization of MSE scores across DNA methylation subtypes, standardized MSE scores were grouped by methylation subtype, and within each subtype, patient columns were ordered using hierarchal clustering with cosine distances and average linkages. MSE scores were transformed using arcsinh transformation and heatmaps were generated using Python *seaborn* package.

### Chromosome 22q loss classifier

A supervised Random Forest Classifier was developed using MSE scores to predict chromosome 22q status (intact vs. loss). Patients in the discovery cohort (334) stratified by DNA methylation subtype and randomly split into training (80%) and held-out test (20%) sets. The same split was used in training the corresponding MIL model. Model hyperparameters were optimized using repeated stratified five-fold cross-validation (five folds, five repeats) with performance assessed using ROC-AUC. Hyperparameters explored included the number of trees (200 or 500), maximum tree depth (5-20), minimum samples required for node splitting and leaf nodes, and bootstrap sampling, with the number of features considered at each split set to the square root of the total number of features. The Gini impurity criterion was used for node splitting. Class imbalance was addressed using balanced class weights during model training. Final model performance was evaluated on the test set using class-specific one-vs-rest ROC AUC values, macro-averaged AUC, confusion matrices, and balanced accuracy. Overall differences in morphological set enrichment scores across methylation subtypes across Chr22q status was assessed using Mann-Whitney U tests, were with p-testing adjusted using the Benjamini-Hochberg (**Fig S11.a-b, e, Table S8**). For visualization of MSE scores across Chr22q status, standardized MSE scores were grouped by methylation subtype, and within each subtype, patient columns were ordered using hierarchal clustering with cosine distances and average linkages. MSE scores were transformed using arcsinh transformation and heatmaps were generated using Python *seaborn* package.

### Chromosome 1p loss classifier

A supervised binary classification model was developed to predict chromosome 1p status (intact vs. loss) using standardized MSE scores. An elastic net-regularized logistic regression classifier and balanced class weights was used to account for class imbalance. Patients in the discovery cohort and CNV labels (333) were stratified by CNV status and randomly split into training (80%) and held-out test (20%) sets. The same split was used in training the corresponding MIL model. The standardization was done within the modeling pipeline to avoid data leakage. Model training and hyperparameter optimization were done using grid search with repeated stratified five-fold cross-validation (five folds, 3 repeats) on the training set. Model performance during optimization was assessed using ROC-AUC. Hyperparameters explored included the inverse regularization strength (C= 0.01-0.1) and the L1/L2 mixing parameter (l1_ratio – 0.1-0.9). The logistic regression model was implemented using the SAGA solver with elastic net regularization and a maximum of 8,000 optimization iterations. The model was developed using Python *scikit-learn* package. The best performing model was refit on the full training data and evaluated on the independent test set using ROC AUC, balanced accuracy, confusion matrices. Kruskal-Wallis tests were done to reveal significant features and a Dunn’s test was used to reveal pairwise differences between methylation subtypes. Overall differences in MSE scores across Chr1p status was assessed using Mann-Whitney U tests, were with p-testing adjusted using the Benjamini-Hochberg and we used Delong’s tests to compare ROC-AUC curves with MIL results. For visualization of MSE scores across Chr1p status, standardized MSE scores were grouped by methylation subtype, and within each subtype, patient columns were ordered using hierarchal clustering with cosine distances and average linkages. MSE scores were transformed using arcsinh transformation and heatmaps were generated using Python *seaborn* package.

### Programming and statistical analysis

All computer programming for clinical and WSI data analysis was done using Python (v3.10.18) and Bash. Deep-learning models were developed using both *Pytorch* (v2.7.1) and *TensorFlow* (v2.15.1). Statistical analysis were performed using the following Python packages: *pingouin* (v0.5.5) for intraclass correlation coefficients, *lifelines* (v0.30.0) for log-rank test, Cox proportional hazards models, Harrell’s concordance index, likelihood ratio test, and Kaplan-Meier estimation; scikit-survival for Brier score computation; *SciPy* (v1.15.2) for Fisher’s exact test, Pearson’s chi-square test, Mann-Whitney U tests, Kruskall-Wallis tests; statsmodels (v0.14.5) for multinomial logistic regression Wald tests, Benjamini-Hochberg false discovery rate correction; and *scikit-posthocs* (v0.11.4) for Dunn’s post hoc testing).

### Illustrations

Multiple figure panels were created with Biorender.com. Most plots were created using seaborn and matplotlib Python libraries and compiled using Inkscape software. Clustering heatmap was generated using *ComplexHeatmap* in R.

## Data Availability

All data produced in the present study are available upon reasonable request to the authors. Processed data is available online at https://drive.google.com/drive/folders/1R0OBOIWIrbp_5tiURGnkwRmc_al5kBWZ?usp=share_link

## Acknowledgements

This work was supported by the US National Institutes of Health National Cancer Institute P50CA221747, U54CA302435, National Institute of Neurological Disorders and Stroke U24NS133949, R01NS117104, R01NS118039, and National Library of Medicine award R01LM013523. The authors thank the Northwestern Nervous System Tumor Bank for providing de-identified clinical and imaging data.

## Ethics statement

All study participants participated voluntarily and provided informed consent. Data sharing was approved through the Northwestern University Institutional Review Board, STU00213676 and Northwestern University Nervous System Tumor Bank STU00095863.

## Author Contributions

M.A.A, C.H., L.A.D.C conceived the research idea. M.A.A developed the MSE methodology, carried out data analysis, and model development. M.A.A, C.H, L.A.D.C wrote the paper. K.M. carried out data collection and digitization. H.C. analyzed molecular data. H.Z. provided expertise in biostatics. M.Y. edited the paper and provided expertise on neurooncology. P.J, J.T.A., C.H provided annotation and model validation. D.R. provided expertise on molecular subtyping. C.H. and L.A.D.C jointly supervised the work.

