## Supplemental Figures for "Morphological set enrichment enables interpretable prognostication and molecular profiling of meningiomas"

Craig M. Horbinski, M.D., Ph.D.

Mayo Clinic Florida

3-162S

4500 San Pablo Rd. S

Jacksonville, FL 32224

Lee A.D. Cooper, Ph.D.

Northwestern University

750 N Lake Shore

Rubloff Building 11-106

Chicago Illinois 60611

**ABSTRACT**

Meningiomas are the most common primary brain tumors and, despite their benign reputation, often behave aggressively. Meningiomas are morphologically heterogeneous, yet the full significance of their histologic diversity is unclear. This is in large part because many features are not readily quantifiable by traditional observer-based light microscopy. Molecular testing improves prognostic stratification, but is not universally accessible. We therefore sought to determine whether an artificial intelligence (AI)-trained program could predict specific genomic and epigenomic patterns in meningiomas, and whether it could extract more prognostic information out of standard hematoxylin and eosin (H&E) histopathology than the current WHO classification. To do this, we developed Morphologic Set Enrichment (MSE), an interpretable computational pathology framework that quantifies statistical enrichment of morphologic patterns, cells, and tissue architecture from H&E whole-slide images. The MSE meningioma histology program was able to accurately predict DNA methylation subtypes and concurrent chromosome 1p/22q losses, in the process identifying specific morphologic patterns associated with key genomic and epigenomic alterations. It also added prognostic value independent of standard clinical and pathological variables. These results demonstrate that AI-based quantitative morphologic profiling can capture clinically and biologically relevant information that redefines risk stratification for meningiomas, incorporating histological information not included in existing grading schemes.

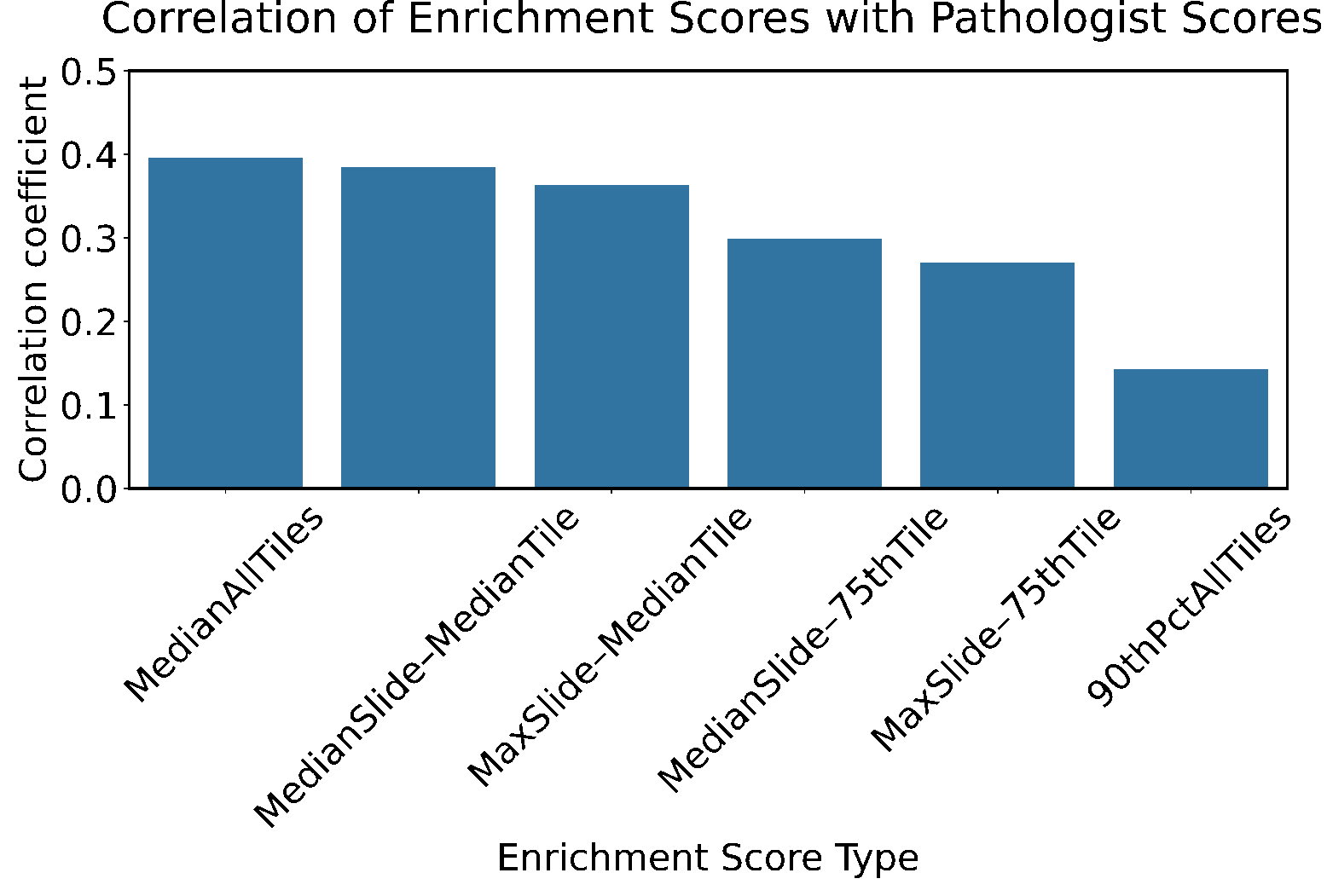

**Figure S1. Patient-level aggregation of patch-level enrichment scores. (a)** Spearman correlations between pathologist’s ordinal score (0-3) on patient-level data and various aggregation methods across patches. The strongest correlation was observed for median of all tiles (median_tiles) and the median of slide-level medians of MSE tile scores (median_medianS).

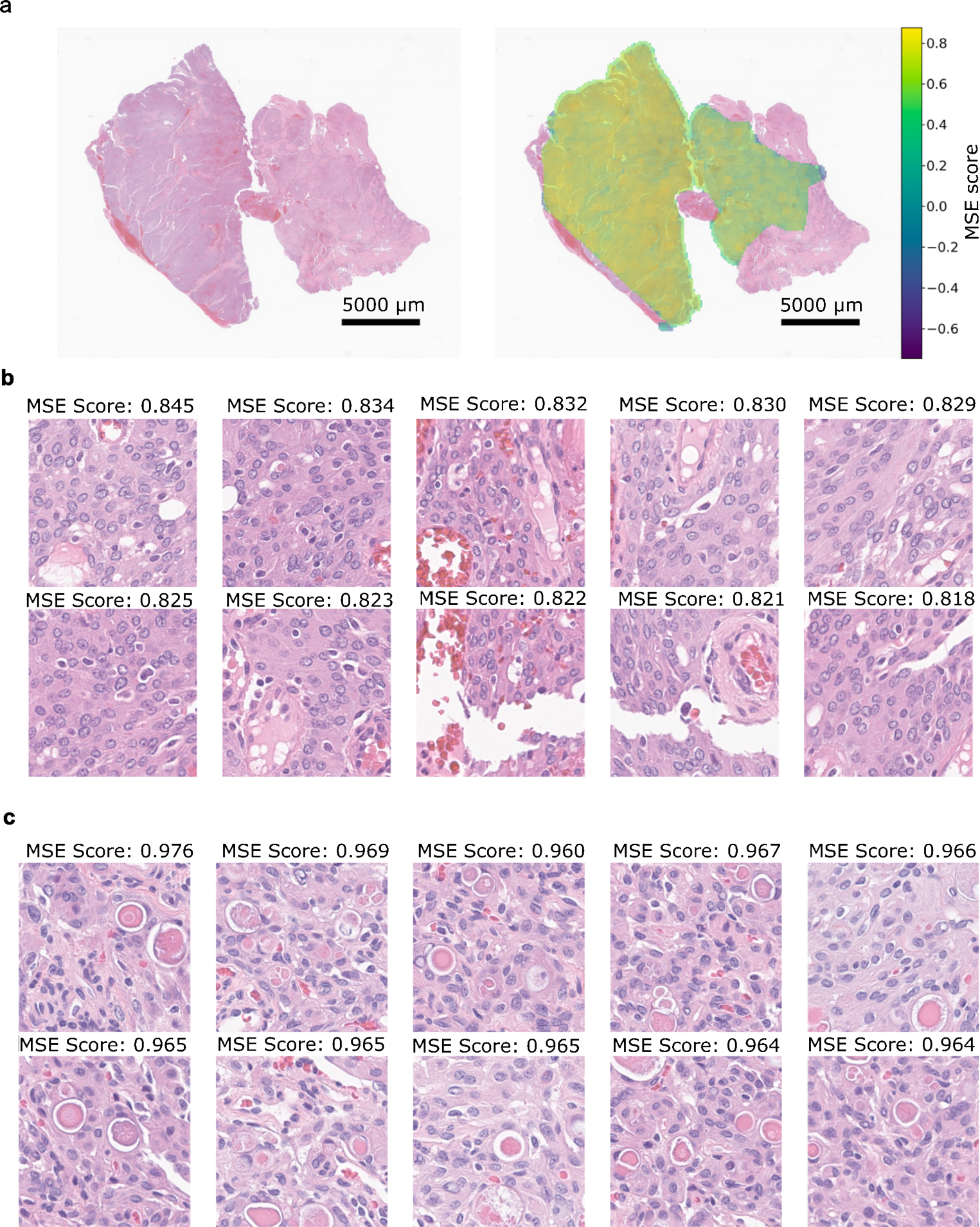

**Figure S2. Validation of secretory samples. (a)** Example slide with high median enrichment score of 0.61 for secretory. (b) Top enrichment scores HPFs with a maximum enrichment score of 0.845. Similarity between image embeddings of blood clusters and serum may contribute to reduced specificity in detection. (c) HPFs from a correctly detected secretory sample (shown in **Fig. 3)** exhibit consistently higher enrichment scores, with a median enrichment score of 0.84 and a maximum of 0.976.

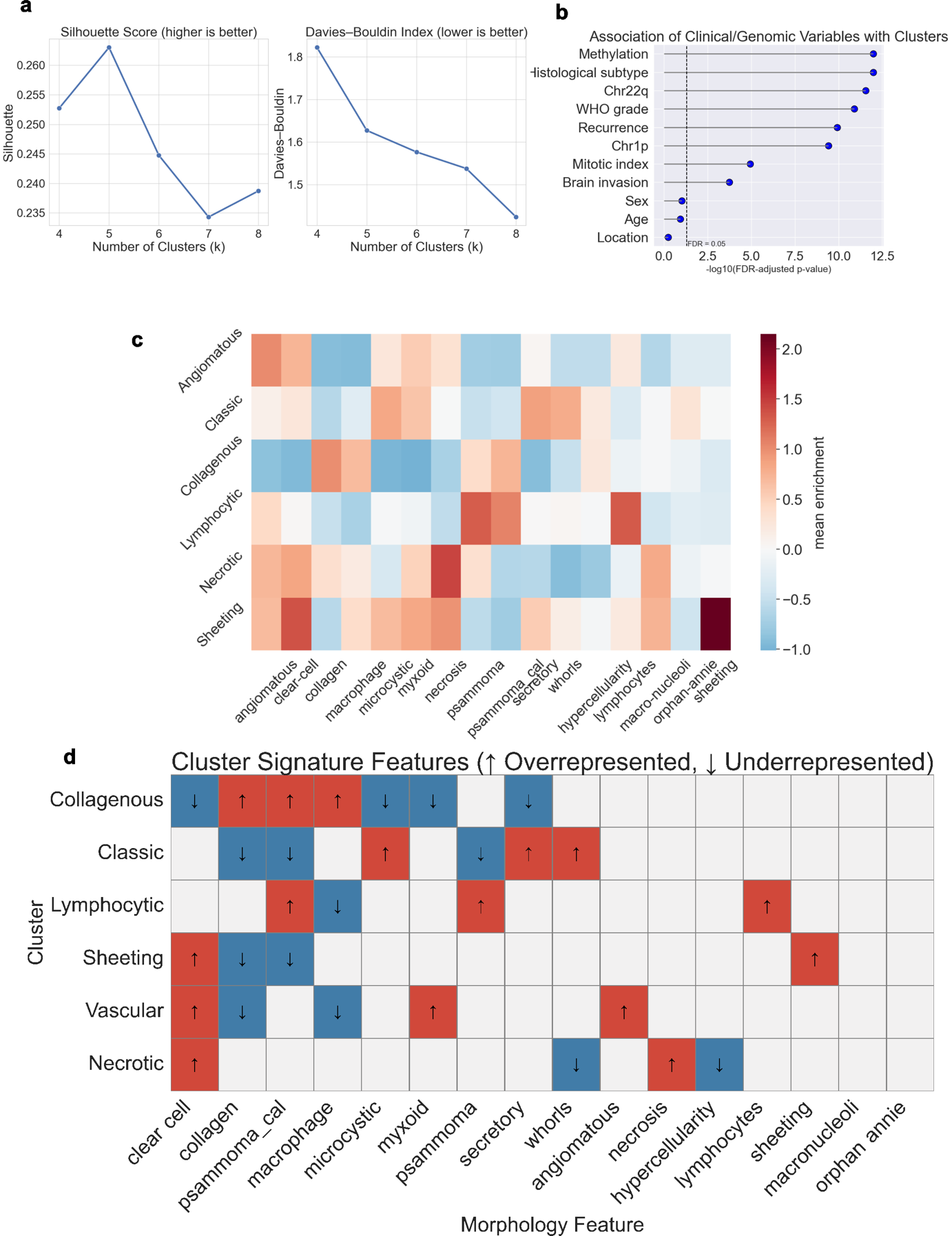

**Figure S3. Clustering MSE scores. (a**) Silhouette and Davies-Bouldin Index values across a range of cluster numbers, identifying 6 clusters as a balance between the two metrics. **(b)** Mean MSE scores within each cluster. (c) Heatmap showing defining combinations of morphologic patterns within clusters. MSE scores higher than 1 standard deviations above the mean were classified as enriched, while those less than 1 of the standard deviations below the mean were classified as depleted.

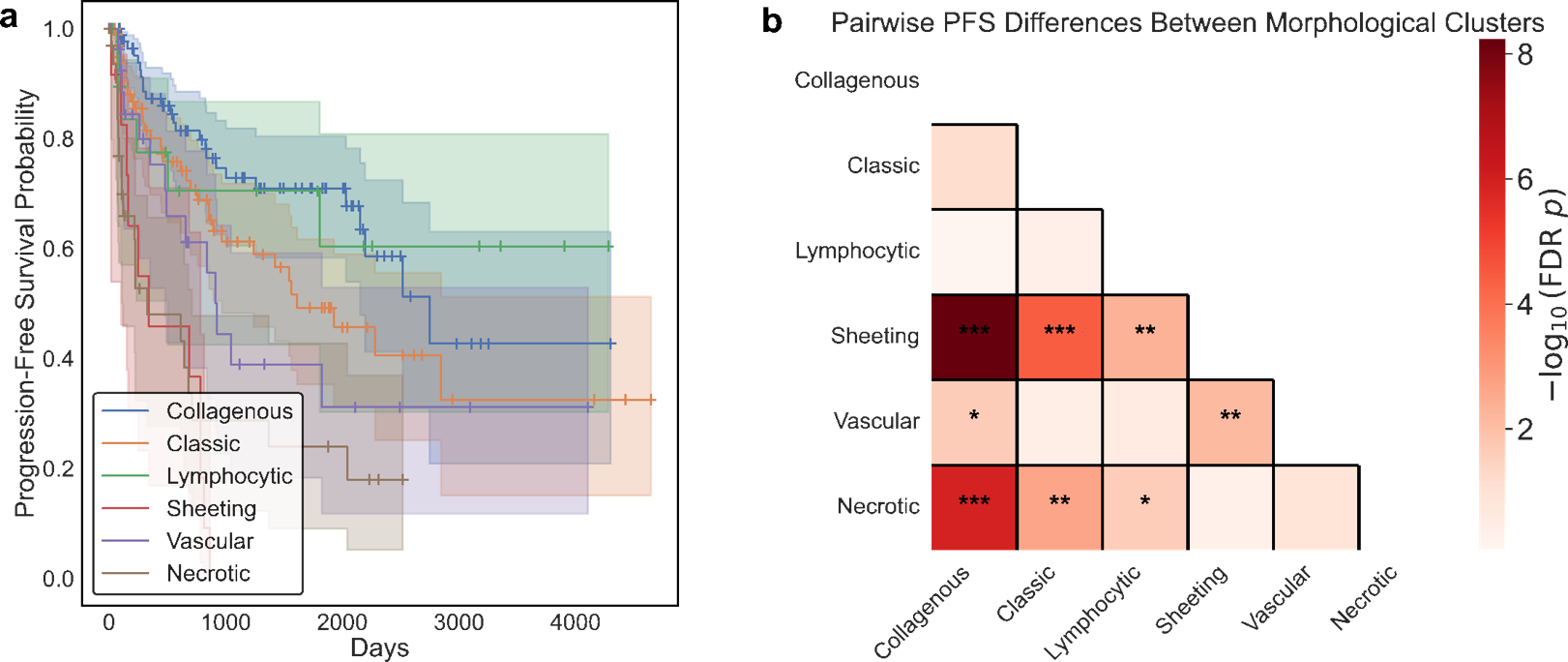

**Figure S4: Cluster rPFS: (a)** Kaplan-Meier curves comparing rPFS among the 6 identified clusters. **(b)** Pairwise log-rank tests evaluating significant survival differences between cluster pairs. (* FDR p-value < 0.05, ** FDR p-value< 0.01, ***FDR p-value< 0.005)**.**

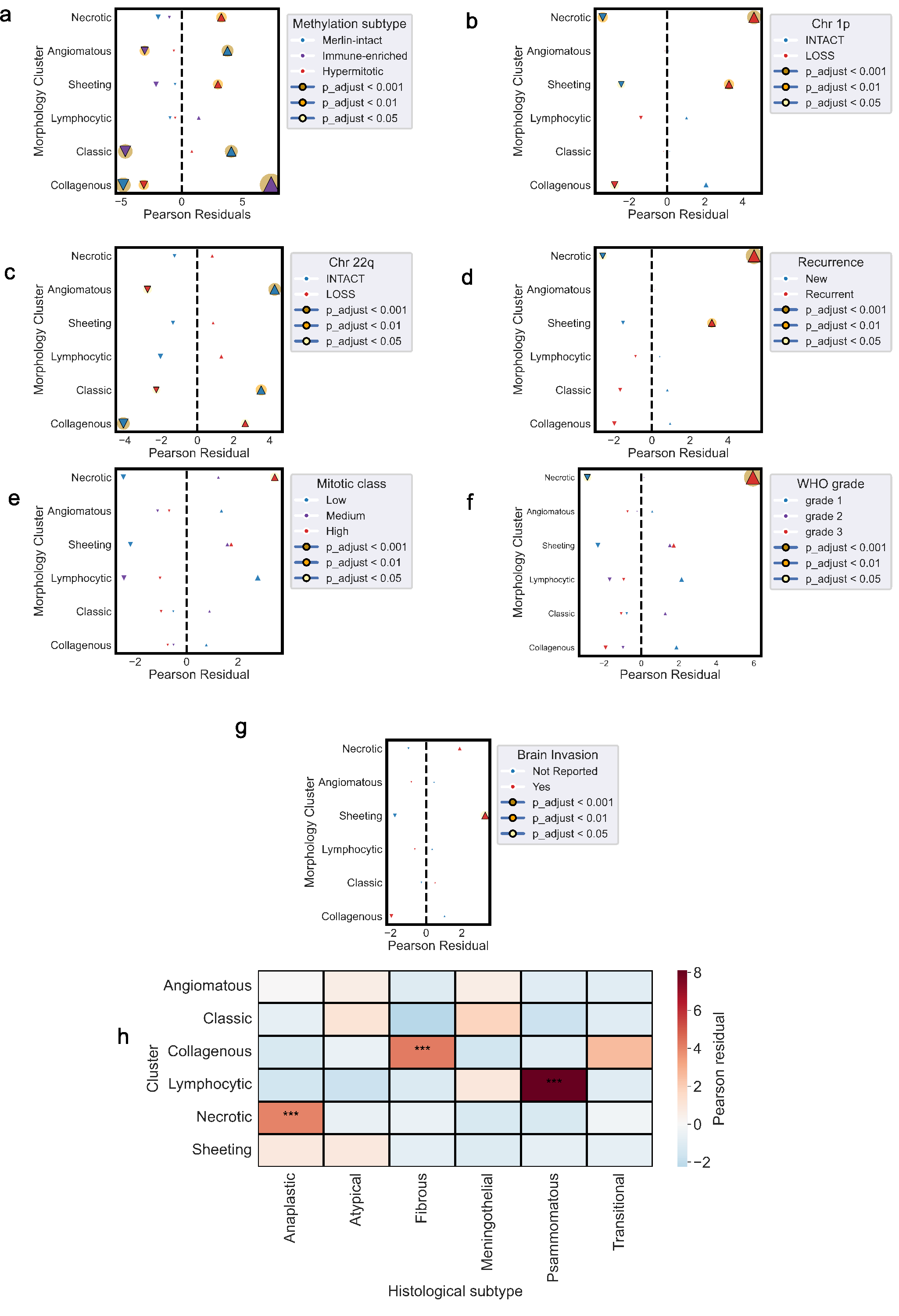

**Figure S5. Clustering associations.** Pearson residuals from chi-square test of independence between morphological clusters and molecular or clinical annotations are shown. Bubble size proportional to -log10(false discovery rate (FDR)-adjusted p-value) and halos denoting significance thresholds. Association are displayed for morphologic patterns and: (**a)** Methylation subtype, (**b)** Chr1p, (**c)** Chr22q, **(d)** Recurrent tumors, **(e)** Mitotic Class, **(f)** WHO grade, **(g)** brain invasion, and **(h)** a heatmap of associations with WHO histological subtypes (* FDR p-value < 0.05, ** FDR p-value< 0.01, ***FDR p-value< 0.005)**.**

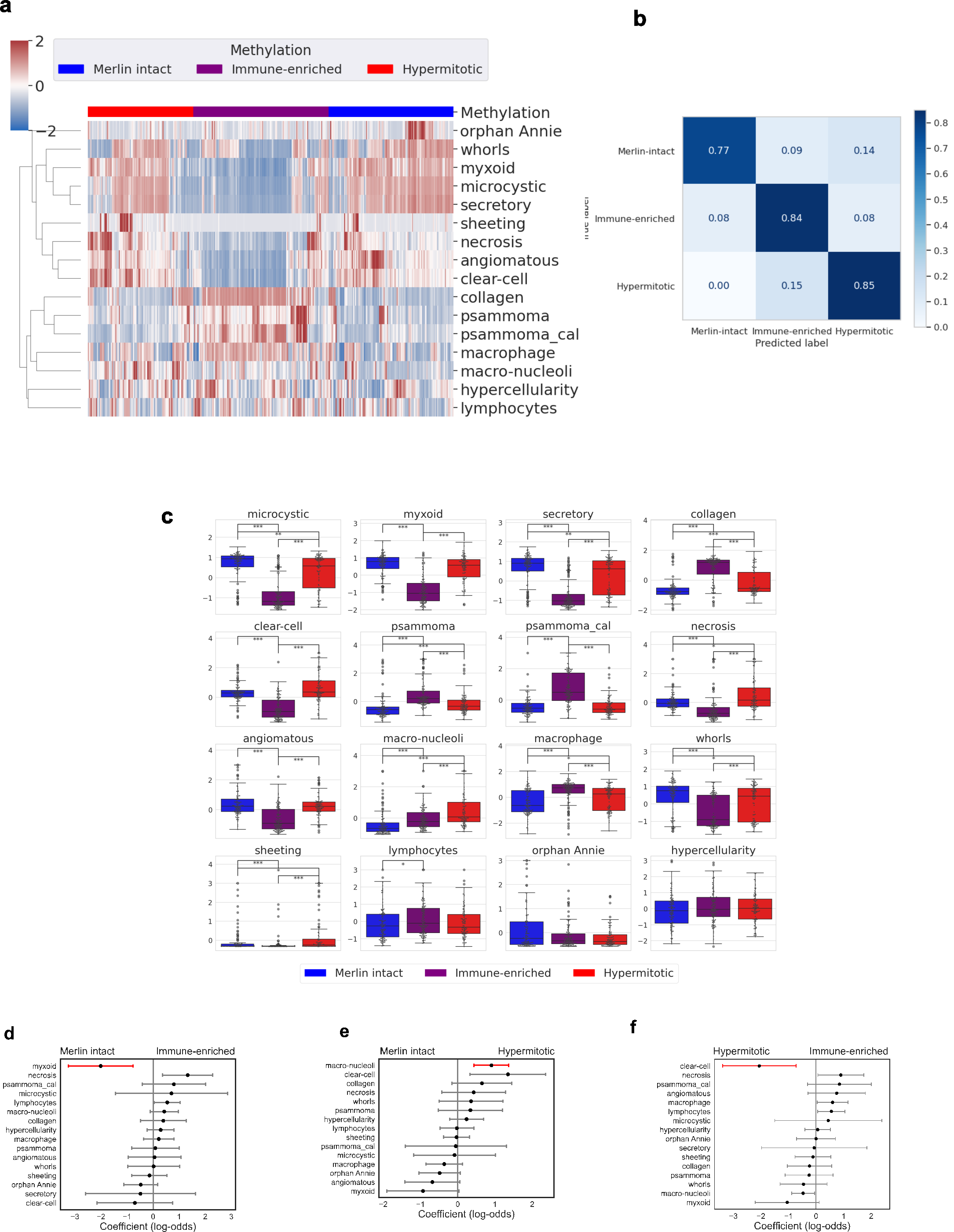

**Figure S6. Morphologic patterns associated with methylation subtypes. (a)** Patients clustered by methylation subtype, revealing distinct morphological associations for each methylation subtype. **(b)** confusion matrix of random forest classifier results for methylation subtypes. **(c)** Box-and-swarm plots of standardized morphological enrichment scores across methylation subtypes, with significant pairwise differences indicated by Dunn’s post hoc tests (Benjamini-Hochberg adjusted). **(f-g)** Morphologic pattern associations with methylation subtypes evaluated by multinomial logistic regression. Red indicates statistically significant patterns (p < 0.05).

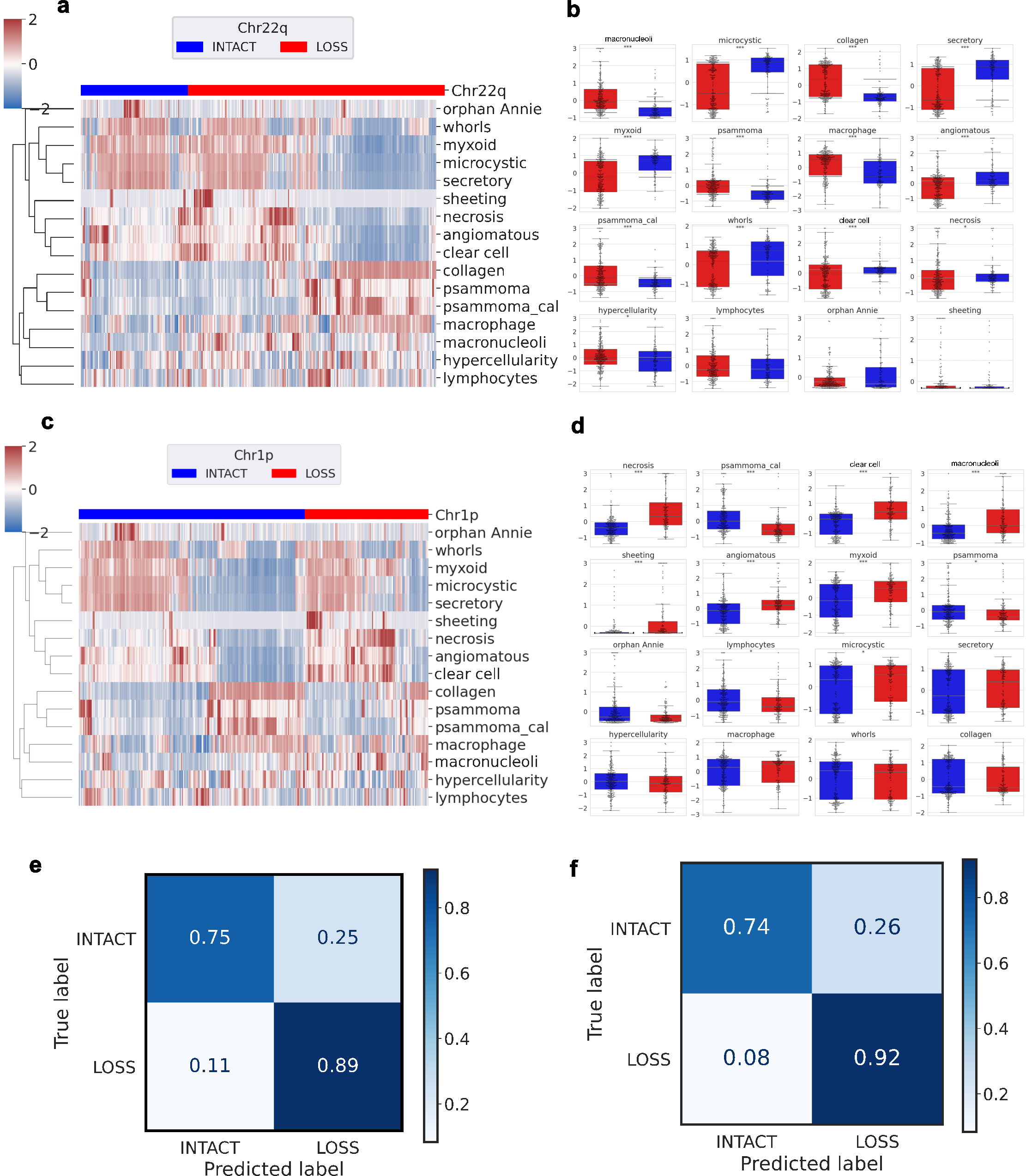

**Figure S7. Morphologic patterns associated with CNV status. (a)** Patients clustered by chromosome 22q status, revealing distinct morphological associations. **(b)** Box-and-swarm plots of standardized morphological enrichment scores across Chr22q status, with significant differences assessed using Mann-Whitney U test (Benjamini-Hochberg adjusted). **(c)** Patients clustered by chromosome 1p status, revealing distinct morphological associations. **(d)** Box-and-swarm plots of standardized morphological enrichment scores across chromosome 1p status, with significant differences assessed using Mann-Whitney U test (Benjamini-Hochberg adjusted). **(e)** Confusion matrix of random forest classifier for chromosome 22q loss. **(f)** Confusion matrix of logistic regression classifier for chromosome 1p loss.

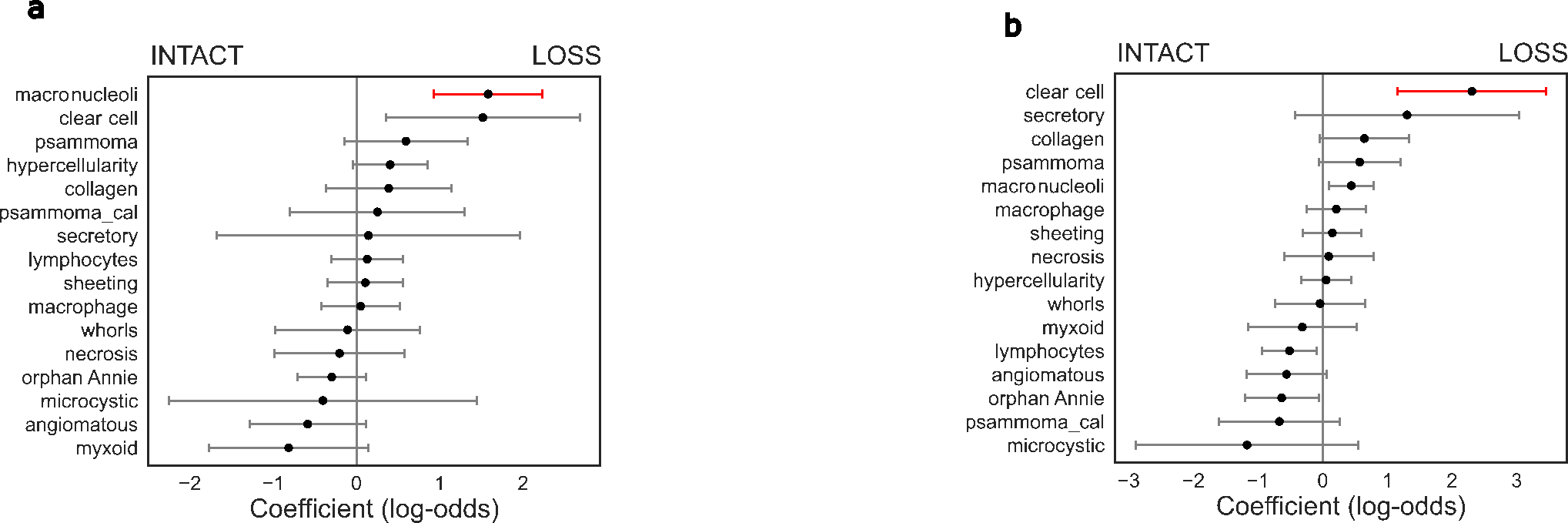

**Figure S8. Multivariable Cox regression for CNVs.** Morphologic pattern associations with **(a)** chromosome 22q, **(b)** chromosome 1p. Red indicates statistically significant morphologic patterns (FDR corrected p < 0.05).

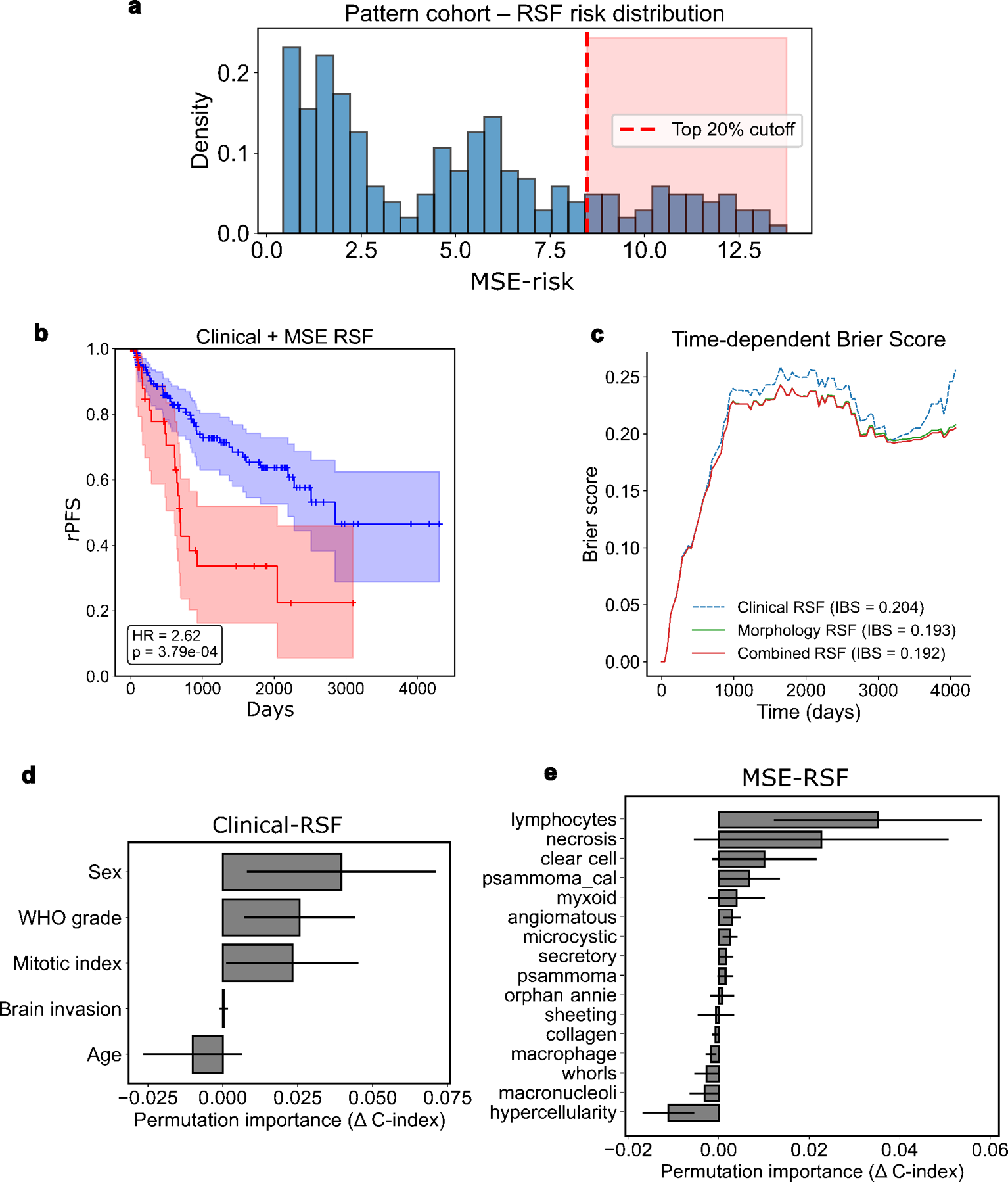

**Figure S9. Random Survival Forests for rPFS prediction (a)** Distribution of MSE-risk in the Pattern cohort, with the high- vs. low-risk cutoff defined at 20^th^ percentile. Cutoff value was subsequently applied to MSE-risk in the discovery cohort. **(b)** Stratification of patients into high- and low-risk groups using the Clinical-MSE-RSF model showing significant difference in rPFS between risk groups. **(c)** Brier scores comparison between Clinical, MSE-RSF, Clinical-MSE-RSF. **(d)** Permutation-based feature importance calculated for Clinical-RSF. **(e)** Permutation-based feature importance calculated for MSE-RSF.

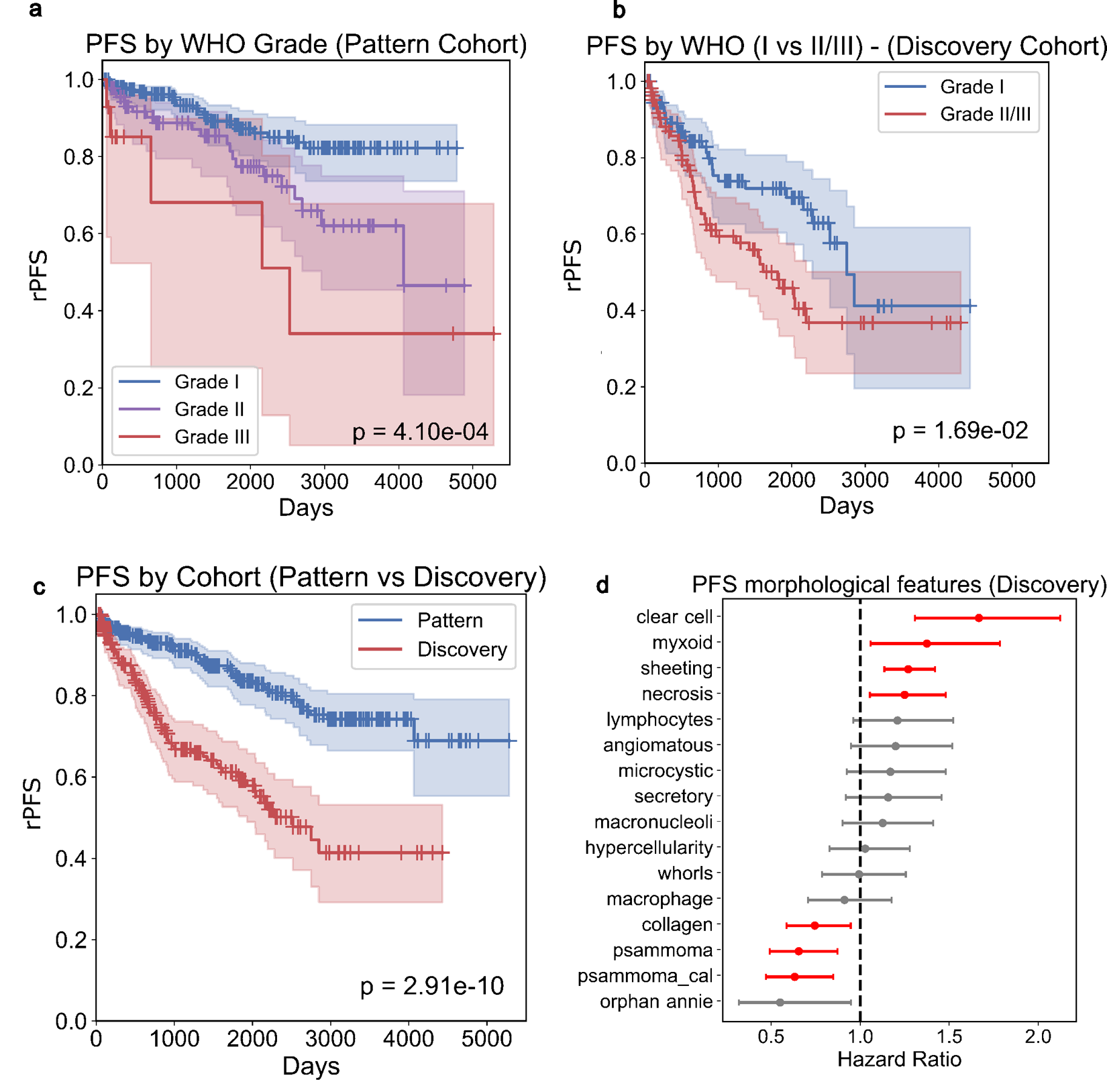

**Figure S10. rPFS analysis:** Kaplan-Meier curves of rPFS stratified by WHO in **(a)**Pattern **(b)**Discovery cohorts. **(c)** Kaplan-Meier plots of rPFS across cohorts, demonstrating a significant difference in survival. **(d)** Univariable Cox regression coefficients of MSE scores in the discovery cohort.

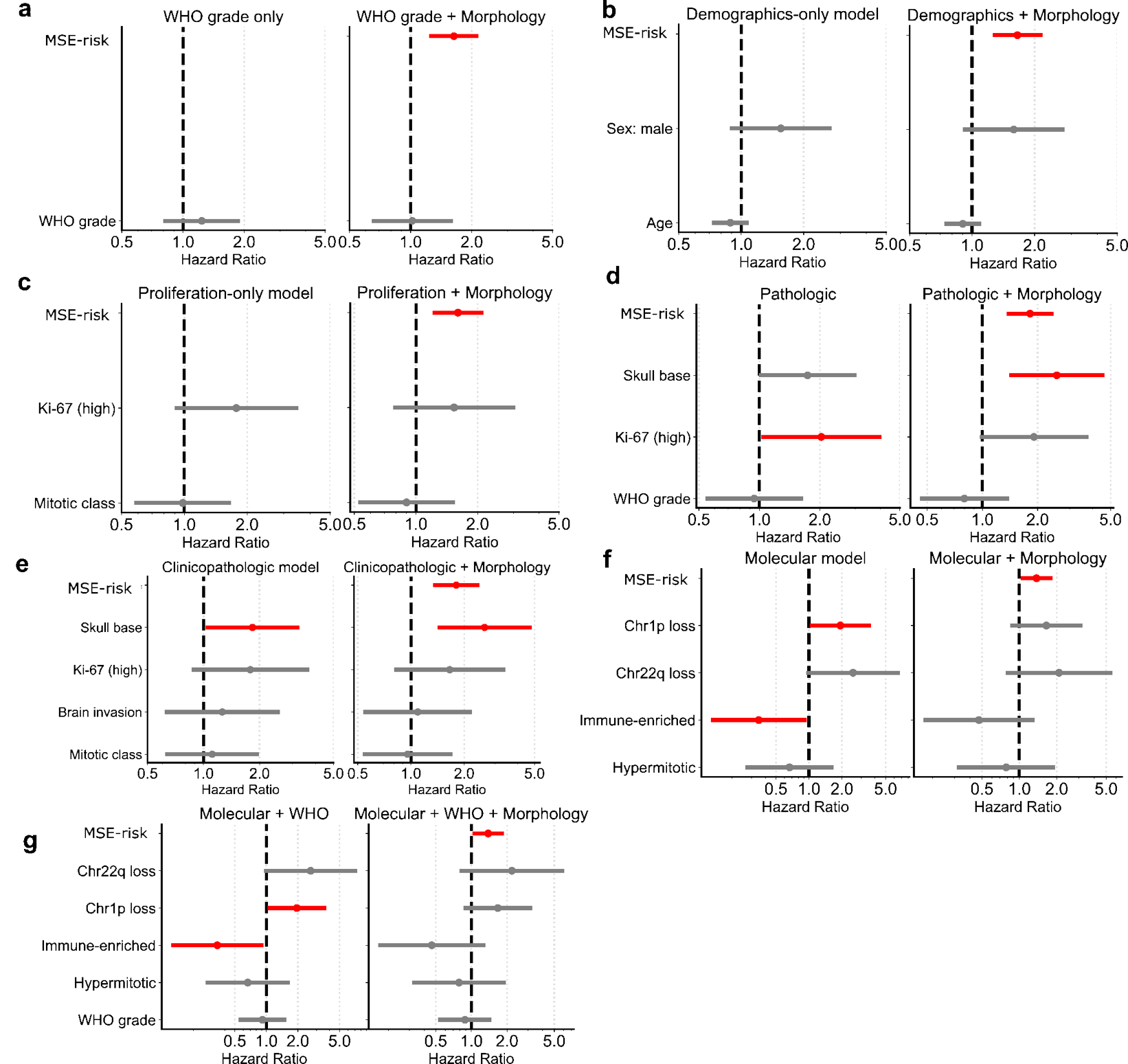

**Figure S11. Independent prognostic value of MSE-risk across covariate sets:** For each covariate set, two multivariable models were fitted: a base model and an extended model incorporating MSE-risk. Covariate groups included **(a)** WHO grade, **(b)** Demographics, **(c)** Proliferation, **(d)** Pathologic, **(e)** Clinicopathologic, **(f)** Molecular biomarkers, **(g)** Molecular and WHO grade. In all cases, adding MSE-risk provided significant independent prognostic value.

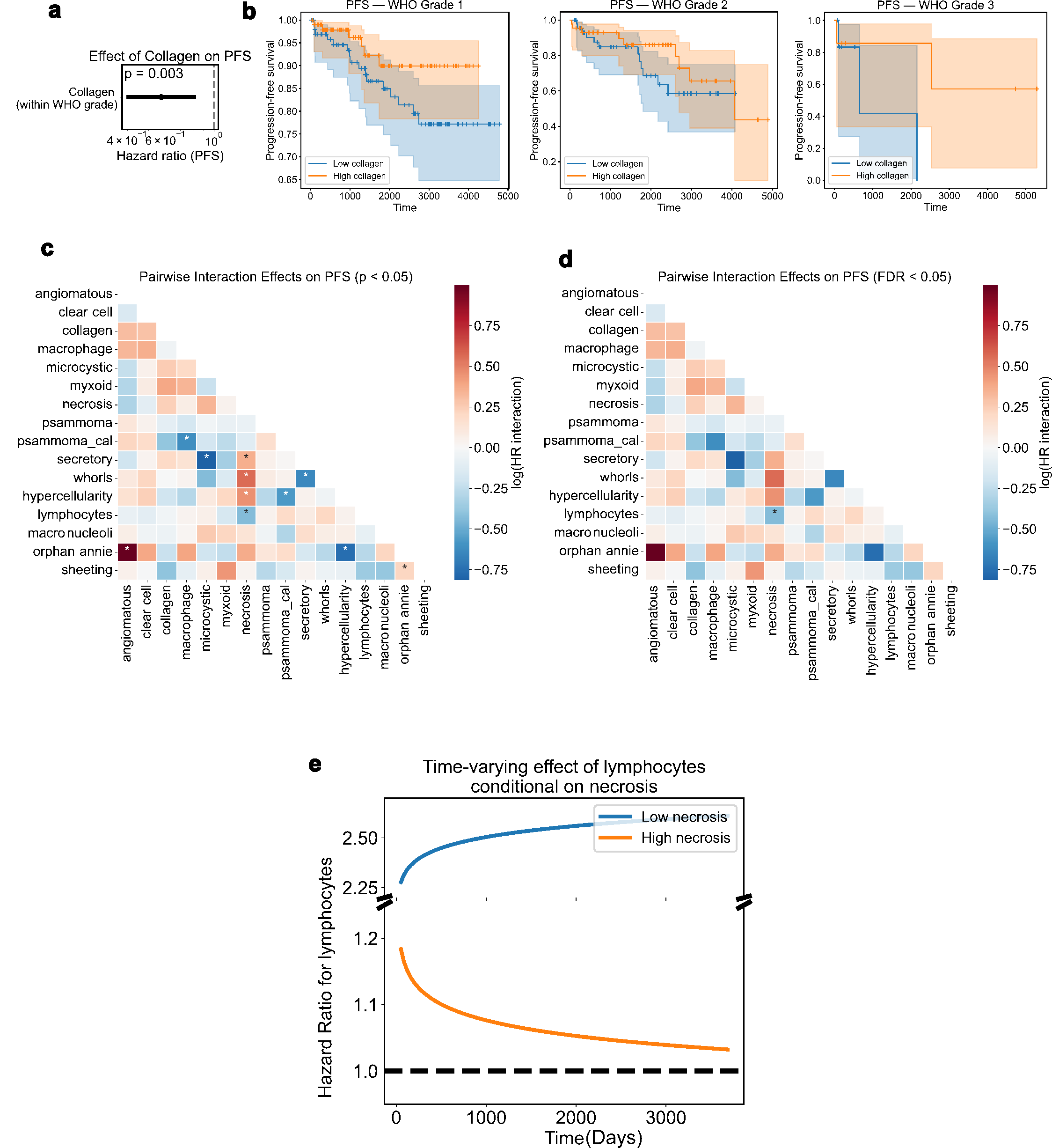

**Figure S12. MSE scores interactions impact of rPFS: (a)** Forest plot showing the effect of collagen on rPFS within WHO grades. Collagen is associated with reduced hazard (p =0.003). **(b)** Kaplan-Meier curves stratified by collagen level where high collagen corresponds to the top 20^th^ percentile of the collagen MSE score. Higher collagen deposition is associated with better rPFS. **(c, d)** Heatmaps of significant pairwise interaction effects among morphologic patterns on rPFS using **(c)** nominal significance (p < 0.05) and **(d)** FDR < 0.05. **(e)** Time-varying hazard ration of lymphocyte infiltration on necrosis MSE score.

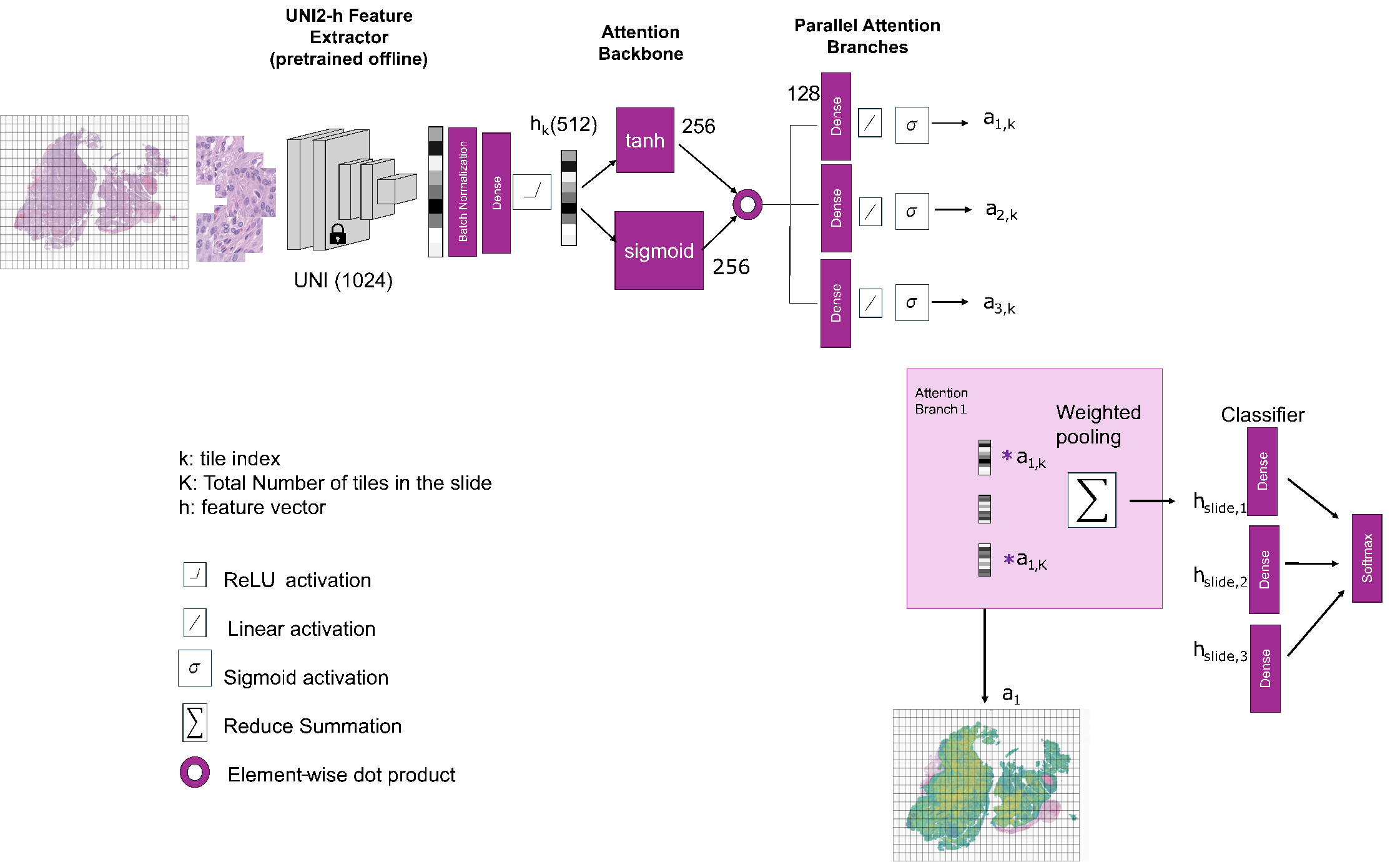

**Figure S13. Multiple instance learning (MIL) model architecture.** Image pixels are inputs to the MIL pipeline. UNI2-h is used for feature extraction at the tile level. For methylation prediction, 3 attention branches are implemented. Weighted pooling aggregated tile-level features, which are then passed to downstream classifiers.
